# Temporal Trends in Racial and Ethnic Disparities in Sleep Duration in the United States, 2004–2018

**DOI:** 10.1101/2021.10.31.21265202

**Authors:** César Caraballo, Shiwani Mahajan, Javier Valero-Elizondo, Daisy Massey, Yuan Lu, Brita Roy, Carley Riley, Amarnath R. Annapureddy, Karthik Murugiah, Johanna Elumn, Khurram Nasir, Marcella Nunez-Smith, Howard P. Forman, Chandra L. Jackson, Jeph Herrin, Harlan M. Krumholz

**Affiliations:** Center for Outcomes Research and Evaluation, Yale New Haven Hospital, New Haven, Connecticut; Section of Cardiovascular Medicine, Department of Internal Medicine, Yale School of Medicine, New Haven, Connecticut; Section of General Internal Medicine, Department of Internal Medicine, Yale School of Medicine, New Haven, Connecticut; Division of Cardiovascular Prevention and Wellness, Houston Methodist DeBakey Heart and Vascular Center, Houston, Texas; Center for Outcomes Research, Houston Methodist Research Institute, Houston, Texas; Department of Chronic Disease Epidemiology, Yale School of Public Health, New Haven, Connecticut; Department of Pediatrics, University of Cincinnati College of Medicine, Cincinnati, Ohio; Division of Critical Care Medicine, Cincinnati Children’s Hospital Medical Center, Cincinnati, Ohio; SEICHE Center for Health and Justice, Section of General Internal Medicine, Yale School of Medicine, New Haven, Connecticut; Equity Research and Innovation Center, Section of General Internal Medicine, Yale School of Medicine, New Haven, Connecticut; Department of Radiology and Biomedical Imaging, Yale School of Medicine, New Haven, Connecticut; Epidemiology Branch, National Institute of Environmental Health Sciences, National Institutes of Health, Department of Health and Human Services, Research Triangle Park, North Carolina; Intramural Program, National Institute on Minority Health and Health Disparities, National Institutes of Health, Department of Health and Human Services, Bethesda, Maryland; Department of Health Policy and Management, Yale School of Public Health, New Haven, Connecticut

## Abstract

**Importance:** Minoritized racial and ethnic groups are generally more likely to experience sleep deficiencies. It is unclear how these sleep duration disparities have changed over recent years.

**Objective:** To determine 15-year trends in the racial and ethnic differences in self-reported sleep duration among adults in the US.

**Design:** Serial cross-sectional study of the National Health Interview Survey from years 2004– 2018. Analyses were performed between July 26, 2021 and February 10, 2022.

**Setting:** US population-based.

**Participants:** 429,195 non-institutionalized adults.

**Exposures:** Self-reported race, ethnicity, household income, and sex/gender.

**Main Outcomes:** Temporal trends and racial/ethnic differences in short- and long-sleep duration (<7 and >9 hours in a 24-hour period, respectively), and racial/ethnic differences in the relationship between sleep duration and age.

**Results:** The study sample consisted of 429,195 individuals (mean age 46.5 [SE, 0.08] years; 51.7% female) of which 5.1% identified as Asian, 11.8% as Black, 14.7% as Latino/Hispanic, and 68.5% as White. In 2004, the adjusted estimated prevalence of short-sleep duration and long-sleep duration, respectively, were 31.3% and 2.5% among Asian individuals, 35.3% and 6.4% among Black individuals, 27.0% and 4.6% among Latino/Hispanic individuals, and 27.8% and 3.5% among White individuals. Over the study period, there was a significant increase in the short sleep prevalence among Black, Latino/Hispanic, and White individuals (P≤0.001 for each), whereas prevalence of long sleep changed significantly only among Latino/Hispanic individuals (−1.4 points, P=0.01). In 2018, compared with White individuals, short sleep prevalence among Black and Latino/Hispanic individuals was higher by 10.7 points and 2.4 points, respectively

(P≤0.03 each), and long sleep prevalence was higher only among Black people (+1.4 points; P=0.01). The short-sleep disparities were the greatest among women and among those with middle/high household income. In addition, across age groups, Black individuals had a higher short- and long-sleep duration prevalence compared with White individuals of their same age.

**Conclusions:** In this serial cross-sectional study from 2004 to 2018, the prevalence of short and long sleep duration was persistently higher among Black individuals. The Black-White disparities in short-sleep were highest among women, individuals who had middle or high income, and among young or middle-aged adults.

**KEY POINTS:** *Question:* How have racial and ethnic differences in self-reported sleep duration among US adults changed between 2004 and 2018?

*Findings:* In this serial cross-sectional study that included 429,195 adults, the prevalence of both short and long sleep duration were persistently higher among Black individuals over the 15-year study period. The disparities in short sleep duration were highest for Black women, Black individuals with middle or high income, and young and middle-aged Black adults.

*Meaning:* There were marked racial and ethnic differences in sleep duration that persisted from 2004 to 2018, which may be contributing to health disparities.

## BACKGROUND

In the US, minoritized racial and ethnic groups are generally more likely to report and experience sleep deficiencies that may be drivers of racial and ethnic disparities in physical health, mental health, and quality of life.^1-9^ Both short and long-sleep duration are more prevalent among Black and Latino/Hispanic people compared with White people.^10-14^ The proportion of people reporting short-sleep duration has increased across different racial and ethnic groups, widening the between Black and White individuals in recent years.^12^ This occurred while a national health objective to increase the proportion of people with sufficient sleep was in place.^15,16^

In 2020, the National Institute on Minority Health and Health Disparities, the National Heart, Lung, and Blood Institute, and the Office of Behavioral and Social Sciences Research proposed a framework for sleep health disparities research that focused on the need for greater understanding of health consequences, and interventions that may eliminate them.^8^ This report underscores the need for a more detailed evaluation of the population-level trends in sleep health disparities.

Several key gaps in knowledge persist. First, though there is information on short sleep, our understanding on trends in disparities in long sleep, also a risk factor for adverse health outcomes, remains poor. Moreover, there is little information on trends in the racial and ethnic disparities in sleep health stratified by age, sex/gender, or household income.^17-19^ For instance, an understanding of how racial and ethnic differences in sleep duration vary with age may illuminate the periods of a lifetime in which these disparities emerge and peak. Furthermore, there are known differences in sleep duration by age and sex/gender,^20^ and people with low income are more likely to report poorer sleep health.^21^ How these differences vary by race and ethnicity remains unknown, and a deeper understanding of these variations is important to identify at-risk groups and institute effective interventions. Finally, many studies have not included Asian people as a distinct, albeit heterogenous, race group.

Accordingly, we describe the temporal trends in racial and ethnic disparities in sleep duration over 15 years using nationally representative data from the National Health Interview Survey (NHIS). We estimated differences in the reported short or long sleep duration between racial and ethnic groups, also stratifying by sex/gender, household income, and health status. In addition, we evaluated the racial and ethnic differences in the relationship between sleep duration and age. The purpose of this study is to illuminate trends in racial/ethnic disparities in sleep duration to inform policies and practices designed to address these disparities.

## METHODS

### Data Source

We used data from the annual NHIS from 2004 to 2018. The NHIS has a complex multistage area probability design that accounts for non-response and oversampling of underrepresented groups, which allows for nationally representative estimates (details in **eMethods**).^22^ We used data from the Sample Adult Core file, which includes responses from one randomly-selected adult from each family for a more in-depth questionnaire (mean conditional and final response rate between 2004-2018 of 80.3% and 62.1%, respectively; **eMethods**). We obtained the harmonized data from the Integrated Public Use Microdata Series Health Surveys (https://nhis.ipums.org/),^23^ including the NHIS strata, primary sampling unit, and person weights. All NHIS respondents provided oral consent prior to participation. The Institutional Review Board at Yale University exempted the study from review as NHIS data are publicly available. The code used to analyze these data is publicly available at https://doi.org/10.5281/zenodo.6028375. The study adheres to the STROBE checklist for reporting crossLsectional studies.

### Study Population

We included individuals ≥18 years old from years 2004 to 2018 of the NHIS. We excluded respondents with missing sleep data. Due to small numbers, we also excluded those who identified as non-Hispanic Alaskan Native/American Indian and those who identified as non-Hispanic and did not select a primary race (details in Results section).

### Demographic Variables

Participants were classified into 4 mutually exclusive subgroups: non-Hispanic Asian (Asian), non-Hispanic Black/African American (Black), Latino/Hispanic, and non-Hispanic White (White), based on their responses to the questions “What race do you consider yourself to be?” and “Do you consider yourself Latino/Hispanic?”. Other characteristics included were self-reported age, sex/gender, household income level, health status, and geographic region. Based on the family income level relative to the respective year’s federal poverty level from the US Census Bureau, income level was categorized as low income (<200%) or middle/high income (≥200%).^24-26^ Other clinical and sociodemographic characteristics were used only to describe the study population (**eMethods**).

### Sleep Duration

In the NHIS, participants were asked “On average, how many hours of sleep do you get in a 24-hour period?” The responses were coded as integers, rounded to the nearest hour (eMethods). We defined recommended sleep duration as 7 to 9 hours of sleep in a 24-hour period, short sleep as reporting sleeping fewer than 7 hours (<7), and long-sleep duration as reporting more than 9 (>9) hours, consistent with expert consensus recommendations.^27^

### Statistical Analysis

We first described the general characteristics of the study population. For each year, we estimated the short and long sleep duration prevalence for each racial/ethnic group using multivariable multinomial logistic regression models, adjusting for age and region (details in **eMethods**). To measure the racial and ethnic differences in short and long sleep duration, we subtracted the annual prevalence among White people from the annual prevalence among Asian, Black, and Latino/Hispanic people for that year. Using these annual estimates and differences between estimates, we determined trends over the study period by fitting weighted linear regression models. In a separate analysis, we tested for an absolute difference in each sleep duration prevalence within each race and ethnic group, and the differences between groups, between 2004 and 2018 using a z-test.

To evaluate the association between race and ethnicity and each of these sleep duration outcomes by age, we used multinomial logistic regression models with categorical sleep duration as the dependent variable and age group as the independent variable (**eMethods**).

We then stratified the main analysis described above by sex/gender and household income, separately. Due to the high amount of missing income information from non-response, the NHIS data include multiply imputed income variables for respondents who do not report income. Thus, our income-stratified analysis was performed based on the National Center for Health Statistics recommendations for multiply imputed data analysis (**eMethods**).^28^ As a supplementary analysis, we also stratified the main temporal trends analysis by health status to explore the extent to which the sleep disparities were explained by racial and ethnic differences in self-perceived health. Finally, we performed a sensitivity analysis to assess if the observed disparities in short-sleep duration between Black and White individuals may be explained alone by differences in self-report bias of sleep duration (eMethods).^29^

For all analyses, a 2-sided P-value <0.05 was used to determine statistical significance. All analyses were performed between July 26, 2021, and February 10, 2022, using Stata SE version 17.0 (StataCorp) and incorporated the NHIS strata, primary sample unit, and sample adult weights to produce nationally representative estimates using the -svy-family of commands for structured survey data. All results are reported with 95% CIs. The NHIS strata, primary sampling unit, and person weights were obtained from IPUMS. All person weights were pooled and divided by the number of years studied, following guidance from the NHIS.^30^

## RESULTS

### Population Characteristics

From 444,743 adults interviewed from 2004 to 2018, we excluded 10,203 (2.3%) who had missing information on sleep duration. Because of small numbers, we also excluded 3440 individuals who identified as non-Hispanic Alaskan Native/American Indian and 1905 individuals who identified as non-Hispanic and did not select a primary race (**eFigure 1**). Thus, the study sample consisted of 429,195 individuals (mean age 46.5 [SE, 0.08] years; 51.7% [95% CI: 51.5, 51.9] female) of which 5.1% (95% CI: 4.9, 5.2) identified as Asian, 11.8% (95% CI: 11.5, 12.2) identified as Black, 14.7% (95% CI: 14.2, 15.1) identified as Latino/Hispanic, and 68.5% (95% CI: 67.9, 69.0) identified as White. Study population characteristics are shown in

**Table 1**, and the unadjusted sleep duration distribution by race and ethnicity is shown in **eFigure 2**.

**Table 1.** Study Population Characteristics.

|  | Asian Individuals | Black Individuals | Latino/Hispanic Individuals | White Individuals | All |
| --- | --- | --- | --- | --- | --- |
| Sample size, n [N=429,195] | 22,924 | 61,226 | 71,567 | 273,478 | 429,195 |
| Age (years), median (IQR) | 42 (31–55) | 42 (29–56) | 38 (28–51) | 48 (33–62) | 46 (31–60) |
| Age category, % (95% CI) |  |  |  |  |  |
| 18–39 years | 44.6 (43.5–45.6) | 44.8 (44.1–45.5) | 53.7 (53.0–54.3) | 34.5 (34.0–34.9) | 39.0 (38.6–39.4) |
| 40–64 years | 42.1 (41.2–43.0) | 42.2 (41.7–42.8) | 37.2 (36.7–37.7) | 44.7 (44.4–45.1) | 43.2 (42.9–43.5) |
| ≥65 years | 13.4 (12.7–14.0) | 12.9 (12.5–13.3) | 9.1 (8.8–9.5) | 20.8 (20.5–21.1) | 17.8 (17.5–18.0) |
| Women, % (95% CI) | 52.8 (52.0–53.6) | 55.0 (54.5–55.6) | 49.3 (48.8–49.8) | 51.6 (51.4–51.8) | 51.7 (51.5–51.9) |
| US Citizenship, % (95% CI) [n=428,343] | 69.7 (68.5–70.8) | 95.3 (94.9–95.6) | 65.2 (64.3–66.1) | 98.5 (98.4–98.5) | 91.8 (91.5–92.0) |
| Education level, % (95% CI) [n=426,934] |  |  |  |  |  |
| Less than high school | 9.4 (8.8–10.1) | 16.8 (16.3–17.3) | 35.3 (34.5–36.0) | 9.5 (9.3–9.7) | 14.1 (13.8–14.4) |
| High school diploma /GED | 26.8 (26.5–27.1) | 30.3 (29.7–30.9) | 26.5 (26.0–27.0) | 26.8 (26.5–27.1) | 26.6 (26.4–26.9) |
| Some college | 22.3 (21.4–23.1) | 33.3 (32.7–34.0) | 25.0 (24.4–25.5) | 31.4 (31.1–31.7) | 30.2 (30.0–30.5) |
| ≥Bachelor's degree | 32.3 (31.8–32.8) | 19.6 (19.0–20.2) | 13.3 (12.8–13.7) | 32.3 (31.8–32.8) | 29.1 (28.6–29.5) |
| Annual income <200% federal poverty level,<sup>a</sup> % (95% CI) | 28.0 (24.9–31.3) | 46.4 (44.1–48.7) | 51.4 (49.4–48.7) | 24.0 (23.0–25.0) | 30.8 (29.9–31.7) |
| Uninsured at the time of interview, % (95% CI) [n=427,762] | 12.0 (11.4–12.6) | 18.1 (17.6–18.6) | 33.4 (32.6–34.2) | 10.3 (10.1–10.6) | 14.7 (14.5–15.0) |
| US region,<sup>b</sup> % (95% CI) |  |  |  |  |  |
| Northeast | 19.7 (18.4–21.2) | 15.9 (15.1–16.8) | 13.7 (12.8–14.6) | 18.9 (18.4–19.5) | 17.8 (17.4–18.3) |
| Midwest | 13.1 (12.0–14.2) | 17.5 (16.6–18.5) | 9.2 (8.3–10.0) | 28.0 (27.3–28.7) | 23.2 (22.7–23.8) |
| South | 22.0 (20.6–23.5) | 58.2 (56.8–59.6) | 36.6 (35.1–38.1) | 33.8 (33.0–34.5) | 36.5 (35.8–37.1) |
| West | 45.2 (43.3–47.1) | 8.4 (7.9–8.9) | 40.6 (39.0–42.2) | 19.4 (18.8–20.0) | 22.5 (22.0–23.1) |
| Married or living with partner, % (95% CI) [n=427,923] | 64.5 (63.5–65.5) | 34.5 (33.9–35.1) | 53.0 (52.4–53.6) | 57.8 (57.3–58.2) | 54.7 (54.3–55.0) |
| Employment status, % (95% CI) [n=428,865] |  |  |  |  |  |

|  | **Asian Individuals** | **Black Individuals** | **Latino/Hispanic Individuals** | **White Individuals** | **All** |
| --- | --- | --- | --- | --- | --- |
| With a job/Working | 65.1 (64.2-66.0) | 59.9 (59.2-60.5) | 65.3 (64.7-65.9) | 62.3 (61.9-62.7) | 62.6 (62.3-62.9) |
| Not in labor force | 30.8 (29.9-31.7) | 31.9 (31.2-32.5) | 29.1 (28.5-29.7) | 34.3 (33.9-34.6) | 33.0 (32.7-33.4) |
| Unemployed | 4.1 (3.8-4.4) | 8.3 (8.0-8.6) | 5.7 (5.4-5.9) | 3.5 (3.4-3.6) | 4.4 (4.3-4.5) |
| Poor or fair health, % (95% CI) | 8.9 (8.4-9.4) | 17.7 (17.3-18.2) | 13.8 (13.4-14.2) | 11.7 (11.5-12.0) | 12.6 (12.4-12.8) |
| Comorbidities, % (95% CI) |  |  |  |  |  |
| Hypertension | 22.2 (21.4-23.0) | 36.2 (35.6-36.9) | 20.7 (20.3-21.2) | 30.4 (30.1-30.7) | 29.3 (29.0-29.5) |
| Diabetes | 7.7 (7.3-8.2) | 11.6 (11.2-11.9) | 9.1 (8.8-9.4) | 8.2 (8.1-8.4) | 8.7 (8.6-8.8) |
| Prior stroke/MI | 2.7 (2.4-3.3) | 5.4 (5.2-5.7) | 3.1 (2.9-3.3) | 6.0 (5.9-6.1) | 5.3 (5.2-5.4) |
| Cancer | 3.1 (2.9-3.4) | 4.2 (4.0-4.4) | 2.9 (2.8-3.1) | 10.6 (10.4-10.7) | 8.3 (8.2-8.4) |
| Emphysema/Chronic bronchitis | 1.8 (1.6-2.0) | 4.6 (4.4-4.8) | 2.8 (2.6-2.9) | 5.8 (5.6-5.9) | 5.0 (4.9-5.1) |
| Current smoker, % (95% CI) | 9.6 (9.1-10.1) | 18.8 (18.3-19.3) | 12.5 (12.2-12.9) | 19.6 (19.3-19.9) | 18.0 (17.8-18.2) |
| Obese (BMI ≥30 kg/m<sup>2</sup>), % (95% CI) | 9.7 (9.2-10.3) | 37.5 (36.9-38.1) | 30.9 (30.-31.4) | 27.0 (26.8-27.3) | 28.0 (27.7-28.2) |
| Abbreviations: BMI, body mass index; CI, confidence interval; GED, general equivalency diploma; IQR, interquartile range; MI, myocardial infarction.<br>Legend: All percentages presented here are weighted percentages.<br>Footnotes:<br><sup>a</sup> Annual household income was categorized relative to the respective year's federal poverty level from the US Census Bureau into middle/high income (≥200%) and low income (<200%). The weighted proportion of individuals with annual income <200% of the federal poverty level was estimated using multiple imputation.<br><sup>b</sup> Based on where the housing unit of the survey participant was located. The 4 regions correspond to the regions recognized by the Census Bureau. | | | | | |

### Temporal Trends over the Study Period from 2004 to 2018

#### Short-Sleep Duration

In 2004, the age- and region-adjusted estimated prevalence of short sleep (<7 hours) was 31.4% among Asian individuals (95% CI: 28.1-34.8), 35.3% among Black individuals (95% CI: 33.4-37.2), 27.0% among Latino/Hispanic individuals (95% CI: 25.4-28.6), and 27.8% among White individuals (95% CI: 27.1-28.6; **Figure 1**). From 2004 to 2018, the prevalence of short sleep significantly increased among Black, Latino/Hispanic, and White people regardless of sex/gender or household income stratum (P≤0.02 for each; **Table 2 and eTable 1**). In the same period, the difference between White and Latino/Hispanic individuals increased significantly (+3.4% [95% CI: 0.6-6.2; P=0.02) but did not change significantly between the other subgroups. In 2018, compared with the estimated prevalence among White individuals (31.0% [95% CI: 30.1-31.9]), short-sleep duration among Black and Latino/Hispanic individuals was higher by 10.7 points (95% CI: 8.1-13.2; P<0.001) and 2.4 points (95% CI: 0.2-4.7; P=0.03), respectively (**Table 2**). The observed Black-White disparities remained in our sensitivity analysis that accounted for differences in overestimation of sleep duration between the two groups (**eFigure 3**).

**Figure 1.**
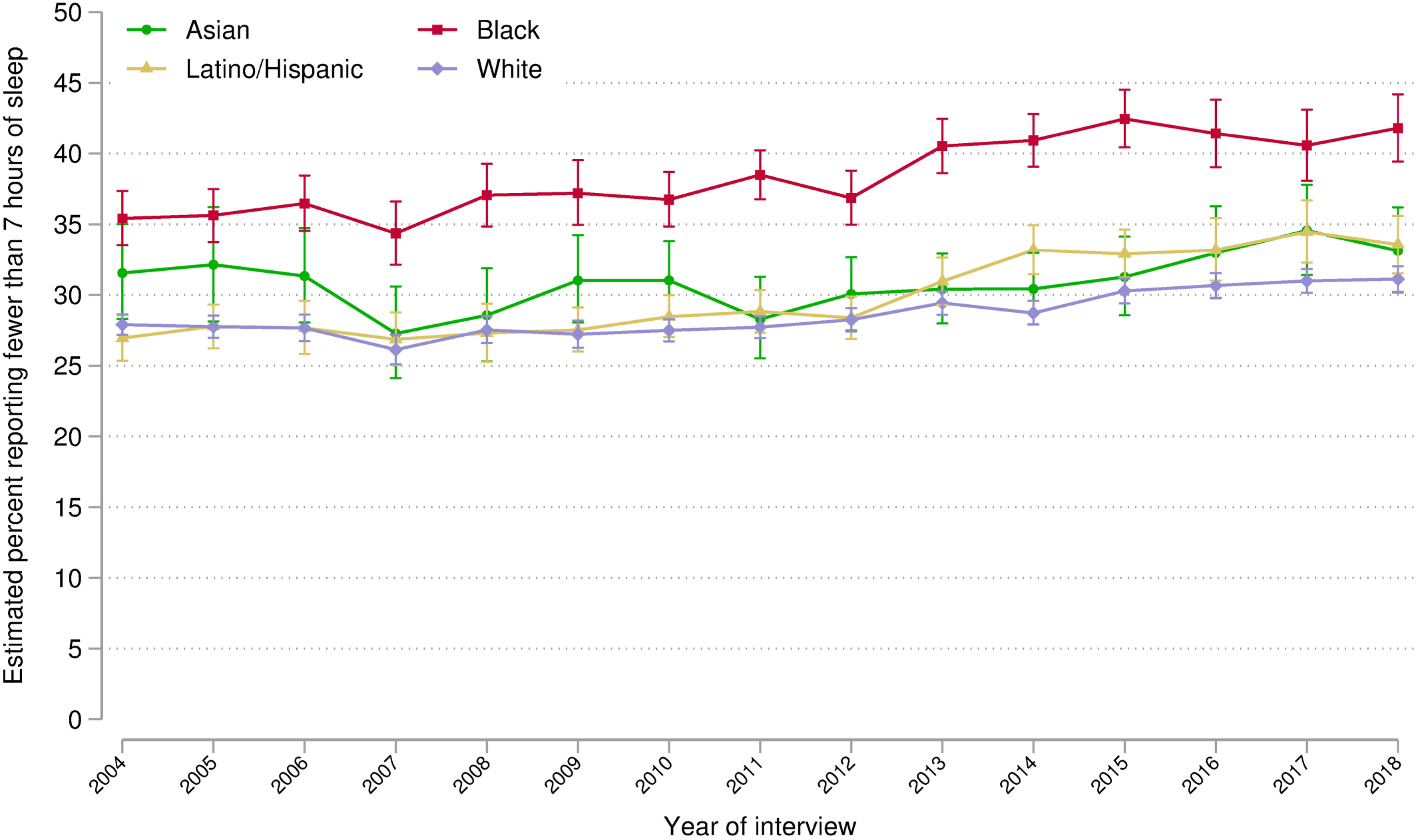
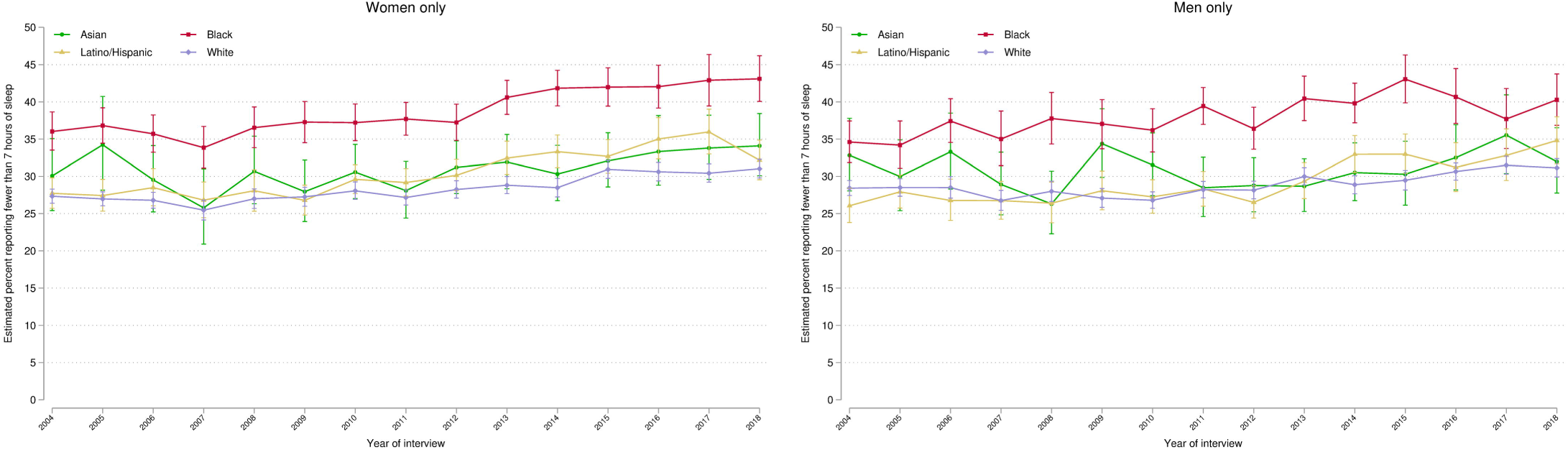
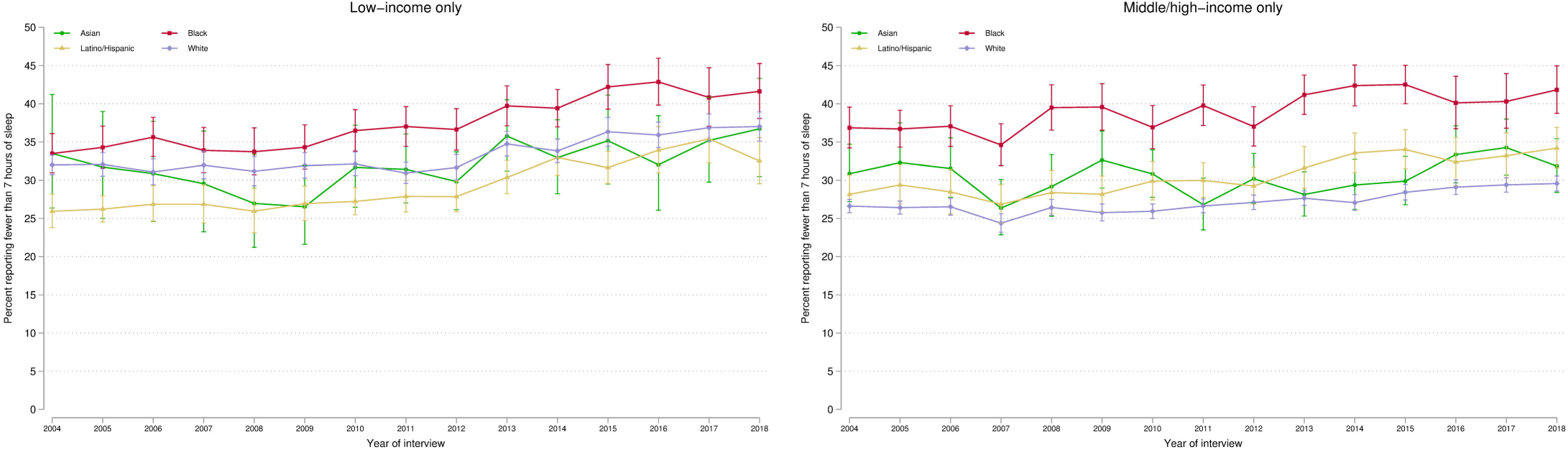
Annual Estimated Prevalence of Short-Sleep Duration by Race and Ethnicity Among US Adults. Data source is the National Health Interview Survey from years 2004 to 2018. Panel A displays overall estimates. Panel B displays estimates stratified by sex/gender. Panel C displays estimates stratified by household income level. Annual prevalence estimates were obtained using multinomial logistic regression adjusted by age and US region (details in Methods and eMethods). Brackets represent 95% confidence intervals. Short-sleep duration was defined as self-reported sleep duration of fewer than 7 (<7) hours in a 24-hour period.

**Table 2.** Change in Short-Sleep Duration and Long-Sleep Duration Prevalence by Race and Ethnicity, 2004 to 2018.

|  | **Asian individuals**<br>Percentage points (95%<br>CI) | **P value** | **Black individuals**<br>Percentage points (95%<br>CI) | **P value** | **Latino/Hispanic**<br>**individuals**<br>Percentage points (95%<br>CI) | **P value** | **White individuals**<br>Percentage points (95%<br>CI) | **P value** |
| --- | --- | --- | --- | --- | --- | --- | --- | --- |
| **Short Sleep (<7 hours)** |  |  |  |  |  |  |  |  |
| *Absolute change in*<br>*prevalence, 2004–2018* |  |  |  |  |  |  |  |  |
| All | +1.58 (-2.95 to 6.10) | 0.50 | +6.39 (3.32 to 9.46) | <0.001 | +6.61 (4.03 to 9.20) | <0.001 | +3.22 (2.06 to 4.38) | <0.001 |
| Women | +4.02 (-2.35 to 10.40) | 0.22 | +7.07 (3.07 to 11.07) | <0.001 | +4.44 (1.10 to 7.77) | 0.009 | +3.69 (2.13 to 5.26) | <0.001 |
| Men | -0.84 (-7.40 to 5.72) | 0.80 | +5.67 (1.20 to 10.14) | 0.01 | +8.75 (4.84 to 12.66) | <0.001 | +2.72 (1.12 to 4.31) | 0.001 |
| Low income | +3.24 (-6.94 to 13.41) | 0.53 | +8.14 (3.57 to 12.72) | <0.001 | +6.57 (2.77 to 10.38) | <0.001 | +5.02 (2.65 to 7.39) | <0.001 |
| Mid/high income | +0.98 (-4.24 to 6.20) | 0.71 | +4.97 (0.77 to 9.18) | 0.02 | +6.04 (2.33 to 9.75) | 0.001 | +2.96 (1.63 to 4.28) | <0.001 |
| *Difference with White,*<br>*2004* |  |  |  |  |  |  |  |  |
| All | +3.65 (0.20 to 7.09) | 0.04 | +7.51 (5.45 to 9.57) | <0.001 | -0.95 (-2.73 to 0.82) | 0.29 | - | - |
| Women | +2.73 (-2.16 to 7.62) | 0.27 | +8.69 (5.97 to 11.41) | <0.001 | +0.40 (-1.84 to 2.63) | 0.73 | - | - |
| Men | +4.40 (-0.59 to 9.39) | 0.08 | +6.18 (3.21 to 9.16) | <0.001 | -2.36 (-4.87 to 0.15) | 0.07 | - | - |
| Low income | +1.49 (-6.30 to 9.28) | 0.71 | +1.49 (-1.59 to 4.56) | 0.34 | -6.06 (-8.69 to -3.42) | <0.001 | - | - |
| Mid/high income | +4.24 (0.33 to 8.16) | 0.03 | +10.23 (7.31 to 13.16) | <0.001 | +1.54 (-1.16 to 4.25) | 0.26 | - | - |
| *Difference with White,*<br>*2018* |  |  |  |  |  |  |  |  |
| All | +2.00 (-1.15 to 5.16) | 0.21 | +10.68 (8.12 to 13.24) | <0.001 | +2.44 (0.23 to 4.65) | 0.03 | - | - |
| Women | +3.06 (-1.31 to 7.43) | 0.17 | +12.07 (8.74 to 15.39) | <0.001 | +1.14 (-1.79 to 4.06) | 0.45 | - | - |
| Men | +0.84 (-3.71 to 5.39) | 0.72 | +9.14 (5.44 to 12.84) | <0.001 | +3.68 (0.28 to 7.07) | 0.03 | - | - |
| Low income | -0.30 (-7.26 to 6.66) | 0.93 | +4.61 (0.47 to 8.74) | 0.03 | -4.51 (-8.14 to -0.88) | 0.02 | - | - |
| Mid/high income | +2.27 (-1.52 to 5.89) | 0.23 | +12.25 (8.95 to 15.55) | <0.001 | +4.62 (1.76 to 7.49) | 0.002 | - | - |
| *Absolute change in*<br>*difference with White,*<br>*2004–2018* |  |  |  |  |  |  |  |  |
| All | -1.64 (-6.32 to 3.03) | 0.49 | +3.17 (-0.11 to 6.46) | 0.06 | +3.39 (0.56 to 6.23) | 0.02 | - | - |
| Women | +0.33 (-6.23 to 6.89) | 0.92 | +3.38 (-0.91 to 7.68) | 0.12 | +0.74 (-2.94 to 4.43) | 0.69 | - | - |
| Men | -3.56 (-10.31 to 3.20) | 0.30 | +2.95 (-1.79 to 7.70) | 0.22 | +6.04 (1.81 to 10.26) | 0.005 | - | - |
| Low income | -1.79 (-12.24 to 8.66) | 0.74 | +3.12 (-2.03 to 8.27) | 0.24 | +1.55 (-2.94 to 6.03) | 0.50 | - | - |
| Mid/high income | -1.97 (-7.36 to 3.41) | 0.47 | +2.02 (-2.39 to 6.43) | 0.37 | +3.08 (-0.86 to 7.02) | 0.13 | - | - |
| **Long Sleep (>9 hours)** |  |  |  |  |  |  |  |  |
| *Absolute change in*<br>*prevalence, 2004–2018* |  |  |  |  |  |  |  |  |
| All | -0.12 (-1.84 to 1.60) | 0.89 | -1.24 (-2.68 to 0.20) | 0.09 | -1.42 (-2.52 to -0.32) | 0.01 | +0.17 (-0.34 to 0.68) | 0.51 |
| Women | -0.81 (-3.42 to 1.80) | 0.54 | -0.51 (-2.32 to 1.30) | 0.58 | -2.05 (-3.46 to -0.64) | 0.004 | +0.15 (-0.43 to 0.74) | 0.61 |
| Men | +0.44 (-1.61 to 2.48) | 0.68 | -2.13 (-4.24 to -0.02) | 0.05 | -0.78 (-2.41 to 0.89) | 0.36 | +0.28 (-0.42 to 0.97) | 0.44 |
| Low income | -2.30 (-6.81 to 2.21) | 0.32 | -2.12 (-4.65 to 0.41) | 0.10 | -1.39 (-3.01 to 0.24) | 0.09 | +0.72 (-0.47 to 1.91) | 0.24 |
| Mid/high income | +0.78 (-0.80 to 2.36) | 0.34 | -0.51 (-2.17 to 1.16) | 0.55 | -1.28 (-2.88 to 0.32) | 0.12 | +0.18 (-0.34 to 0.69) | 0.50 |
| *Difference with White,*<br>*2004* |  |  |  |  |  |  |  |  |
| All | -0.92 (-2.41 to 0.56) | 0.22 | +2.90 (1.81 to 3.99) | <0.001 | +1.09 (0.33 to 1.85) | 0.005 | - | - |
| Women | -0.11 (-2.35 to 2.13) | 0.92 | +2.49 (1.17 to 3.80) | <0.001 | +1.54 (0.46 to 2.61) | 0.005 | - | - |
| Men | -1.72 (-3.48 to 0.04) | 0.06 | +3.41 (1.69 to 5.12) | <0.001 | +0.66 (-0.46 to 1.79) | 0.25 | - | - |
| Low income | -0.06 (-4.20 to 4.08) | 0.98 | +3.49 (1.52 to 5.45) | <0.001 | -0.34 (-1.57 to 0.90) | 0.59 | - | - |
| Mid/high income | -1.40 (-2.61 to -0.18) | 0.02 | +1.47 (0.26 to 2.68) | 0.02 | +1.03 (-0.14 to 2.21) | 0.09 | - | - |
| *Difference with White,*<br>*2018* |  |  |  |  |  |  |  |  |
| All | -1.27 (-2.25 to -0.29) | 0.01 | +1.44 (0.39 to 2.48) | 0.007 | -0.55 (-1.47 to 0.36) | 0.24 | - | - |
| Women | -1.07 (-2.55 to 0.40) | 0.15 | +1.83 (0.45 to 3.20) | 0.009 | -0.67 (-1.74, 0.41) | 0.23 | - | - |
| Men | -1.56 (-2.81 to -0.30) | 0.02 | +1.00 (-0.41 to 2.42) | 0.16 | -0.39 (-1.81 to 1.03) | 0.59 | - | - |
| Low income | -3.08 (-5.23 to -0.94) | 0.005 | +0.65 (-1.34 to 2.64) | 0.52 | -2.45 (-4.04 to -0.85) | 0.003 | - | - |
| Mid/high income | -0.79 (-1.93 to 0.35) | 0.17 | +0.79 (-0.46 to 2.05) | 0.22 | -0.42 (-1.62 to 0.78) | 0.49 | - | - |
| *Absolute change in*<br>*difference with White,*<br>*2004–2018* |  |  |  |  |  |  |  |  |
| All | -0.34 (-2.12 to 1.43) | 0.70 | -1.46 (-2.98 to 0.05) | 0.06 | -1.65 (-2.84 to -0.45) | 0.007 | - | - |
| Women | -0.96 (-3.64 to 1.71) | 0.48 | -0.66 (-2.56 to 1.24) | 0.50 | -2.20 (-3.72 to -0.68) | 0.005 | - | - |
| Men | +0.16 (-2.00 to 2.32) | 0.88 | -2.40 (-4.63 to -0.18) | 0.03 | -1.05 (-2.86 to 0.76) | 0.25 | - | - |
| Low income | -3.02 (-7.68 to 1.64) | 0.20 | -2.84 (-5.64 to -0.04) | 0.05 | -2.11 (-4.12 to -0.10) | 0.04 | - | - |
| Mid/high income | +0.61 (-1.06 to 2.27) | 0.47 | -0.68 (-2.42 to 1.07) | 0.47 | -1.45 (-3.13 to 0.23) | 0.09 | - | - |
| Data source is the National Health Interview Survey from years 2004 to 2018. Short-sleep duration was defined as fewer than 7 (<7) hours of sleep in a 24-hour period. Long-sleep duration was defined as more than 9 (>9) hours of sleep in a 24-hour period. For change in prevalence and change in difference: a positive sign (+) means the prevalence (or its difference with White individuals) increased and a negative sign (-) means it decreased. Prevalence estimates were adjusted by age and region. |  |  |  |  |  |  |  |  |
| Abbreviations: CI, confidence interval |  |  |  |  |  |  |  |  |

Similarly, the prevalence difference between Black women and White women persisted during the study period and was +12.1 points (95% CI: 8.7-15.4; P<0.001) in 2018; among men in 2018, the Black-White gap was +9.1 points (95% CI: 5.4-12.8; P<0.001). The prevalence difference between Latino/Hispanic men and White men, absent in 2004, increased and reached +3.7 points (95% CI: 0.3-7.1; P=0.03) in 2018, whereas there was no significant change in the difference between women (**Figure 1 and Table 2**).

When stratified by income, there were no significant changes in the differences between groups over the study period. In 2018, the difference between Black individuals and White individuals in 2018 was +12.3 percentage points (95% CI: 9.0–15.6; P<0.001) among those with middle/high income and +4.6 points (95% CI: 0.5–8.7; P=0.03) among those with low income. In the same year, the difference between Latino/Hispanic individuals and White individuals was +4.6 points (95% CI: 1.8–7.5; P=0.002) and -4.5 points (95% CI: -8.1 to -0.9; P=0.02) among those with middle/high income and low-income, respectively (**Table 2**). The differences in 2018 between White and Asian individuals were not significant, regardless of income level.

#### Long-Sleep Duration

In 2004, the adjusted estimated prevalence of long sleep (>9 hours) was 2.5% among Asian individuals (95% CI: 1.4-4.3), 6.4% among Black individuals (95% CI: 5.4-7.5), 4.6% among Latino/Hispanic individuals (95% CI: 3.9-5.3), and 3.5% among White individuals (95% CI: 3.2-3.8; **Figure 2**). From 2004 to 2018, the prevalence of long sleep significantly changed only among Latino/Hispanic people (−1.4 percentage points [95% CI: -2.5 to -0.3; P=0.01]; **Table 2**). In 2018, compared with the estimated long-sleep prevalence among White individuals (3.7% [95% CI: 3.4-4.1]), prevalence was higher by 1.4 points among Black individuals (95% CI: 0.4-2.5; P=0.007). Compared with White women, Black women had higher prevalence of long sleep during the study period (**Table 2 and Figure 2**). When stratified by income, the 2018 difference between White and Black individuals was not significant (**Figure 2**).

**Figure 2.**
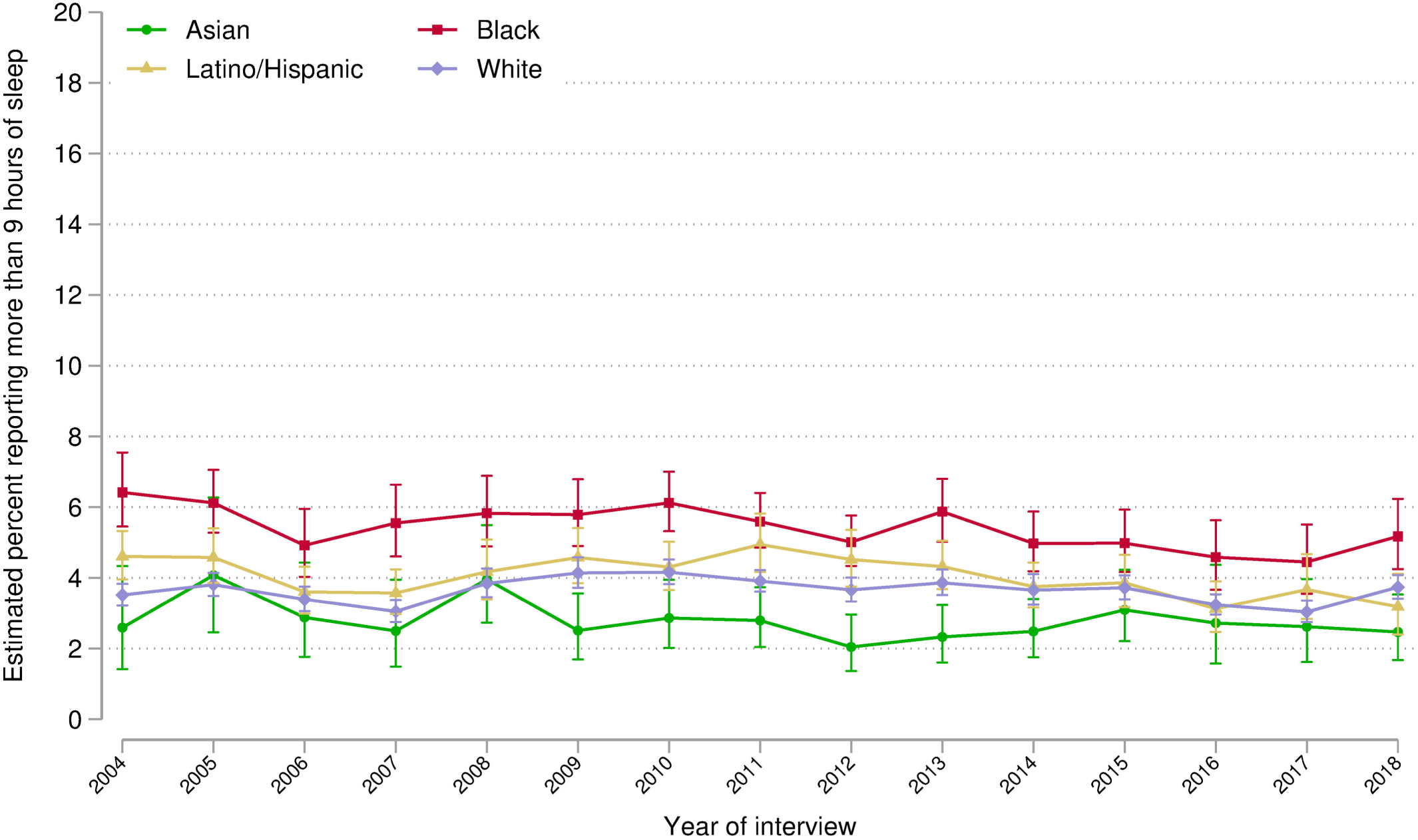
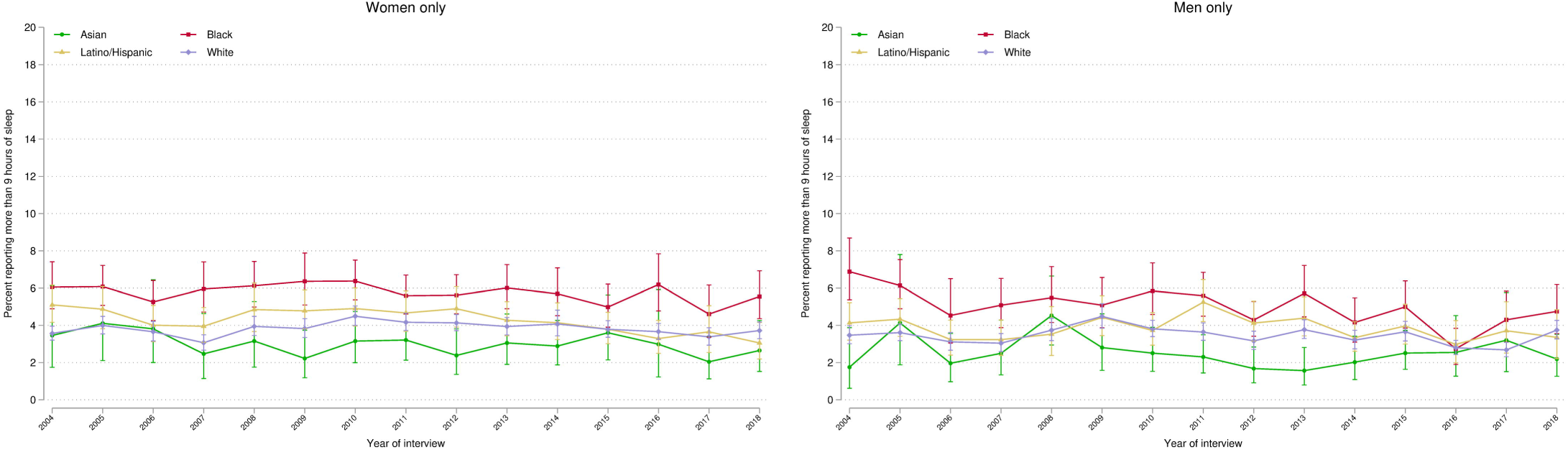
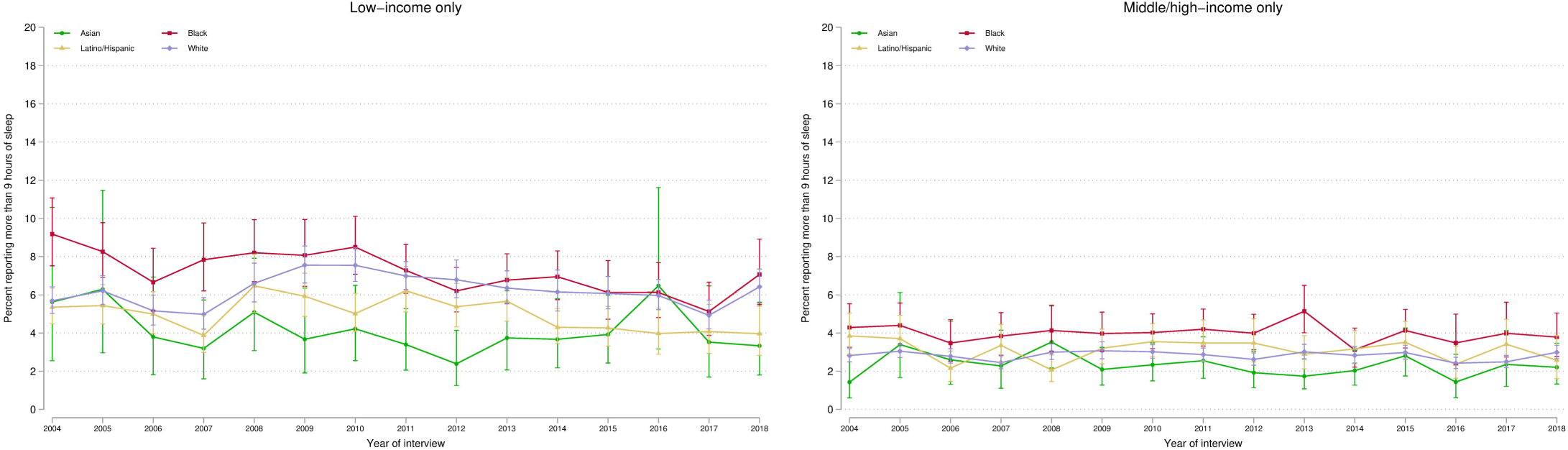
Annual Estimated Prevalence of Long-Sleep Duration by Race and Ethnicity Among US Adults. Data source is the National Health Interview Survey from years 2004 to 2018. Panel A displays overall estimates. Panel B displays estimates stratified by sex/gender. Panel C displays estimates stratified by household income level. Annual prevalence estimates were obtained using multinomial logistic regression adjusted by age and US region (details in Methods and eMethods). Brackets represent 95% confidence intervals. Long-sleep duration was defined as self-reported sleep duration of more than 9 hours (>9) in a 24-hour period.

#### Supplementary Analysis: Stratification by Health Status

When stratified by health status, the observed racial and ethnic disparities in short and long sleep duration persisted within each health status stratum over the study period (**eTables 2 and 3** and **eFigures 4 and 5**).

### Differences in the Relationship Between Sleep Duration and Age

#### Short-Sleep Duration

When compared with White individuals of the same age, short sleep duration was more prevalent among Black people, with a difference starting at +6.9 points among those 18–24 years (95% CI: 5.3-8.5), peaking at +10.7 points among those aged 50–59 years (95% CI: 8.9-12.6), and reaching at 2.9% among those older than 84 years old (95% CI: 0.7-5.1) (**Figure 3 and eFigure 6**). Among those older than 65 years, the prevalence of short-sleep duration decreased for all subgroups as age increased. Similar patterns were observed by sex/gender and among those with middle/high income (**eFigure 7**).

**Figure 3.**
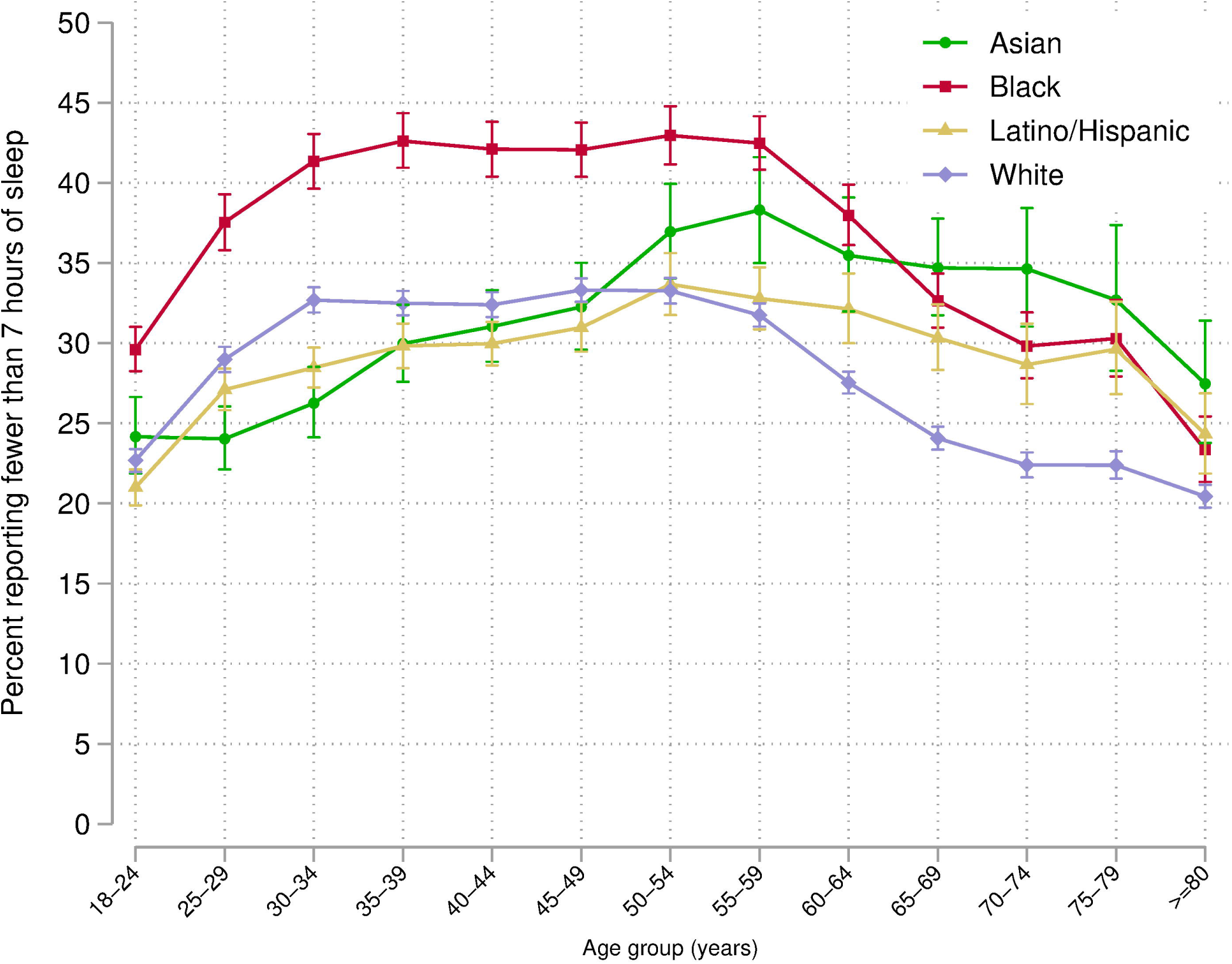
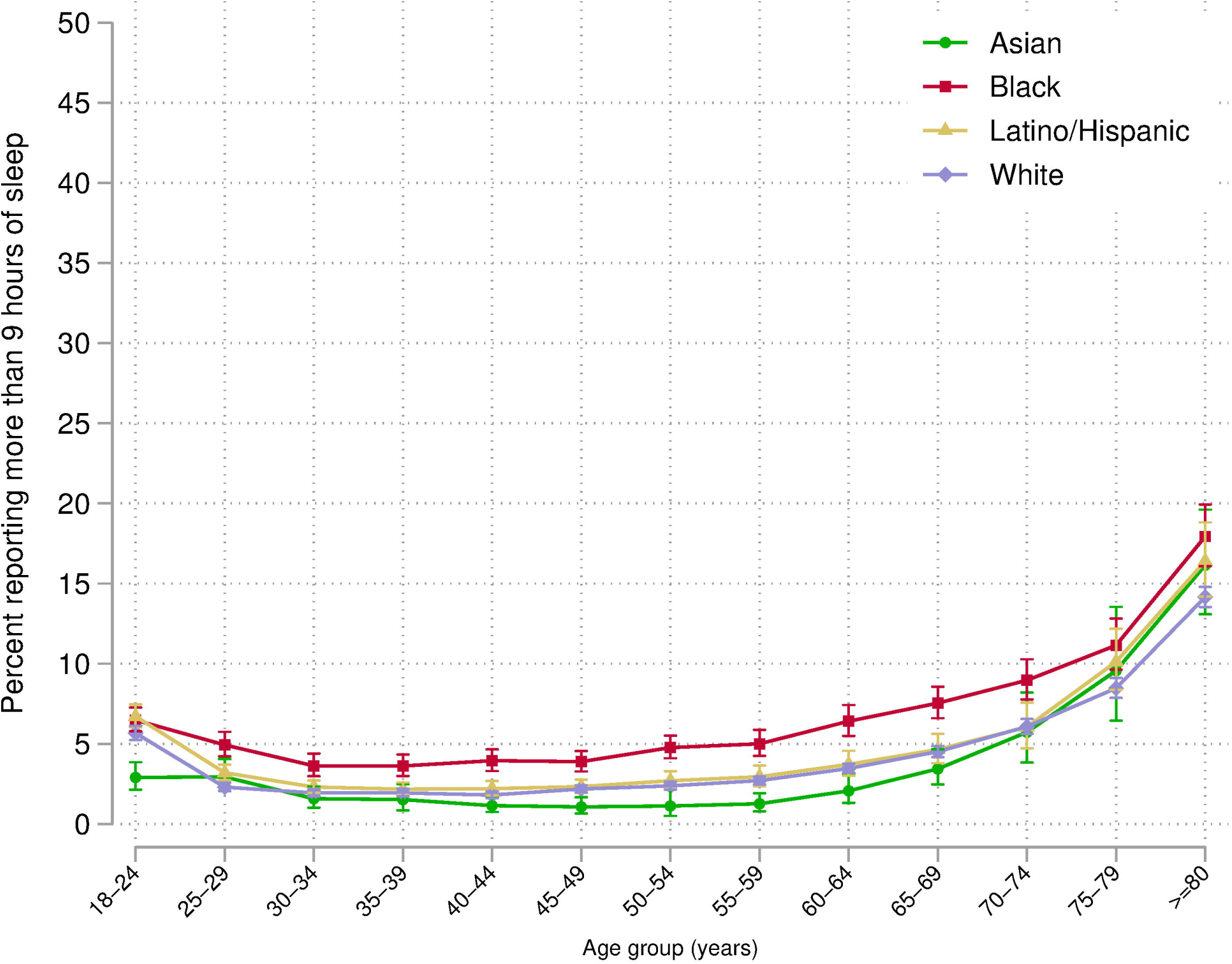
Relationship Between Age and Short- and Long-Sleep Duration by Race/Ethnicity Among US Adults. Data source is the National Health Interview Survey from years 2004 to 2018. Brackets represent 95% confidence intervals. Short-sleep duration was defined as self-reported sleep duration of fewer than 7 (<7) hours in a 24-hour period. Long-sleep duration was defined as self-reported sleep duration of more than 9 (>9) hours in a 24-hour period. Prevalence estimates for each age group were obtained using multinomial logistic regression (details in Methods and eMethods).

#### Long-Sleep Duration

Across all race/ethnicity groups, prevalence of long sleep was lower among those between 30 to 60 years old. Except for individuals aged 18–24 years, Black individuals had a higher prevalence than White individuals across all age groups, ranging from +1.7 points among those aged 30–35 years (95% CI: 0.9-2.4) to +3.8 points among those aged ≥80 years (95% CI: 1.7-5.8; **Figure 3**). Similar patterns were observed by sex/gender and income strata (**eFigure 8**).

## DISCUSSION

In this nationally representative sample from 2004 to 2018, there was increasing prevalence of short sleep duration with persistence of racial and ethnic differences. Black individuals consistently had the highest prevalence of short sleep duration, reaching a difference of 11 percent points compared with White individuals in 2018. The disparities were larger for Black women and Black individuals with middle/high income. In addition, the proportion of Latino/Hispanic individuals who reported short sleep increased among men, widening their gap with White men to nearly 4 percentage points in 2018. Furthermore, Black individuals also had the highest prevalence of long sleep duration over the study period, although this disparity was narrower than that of short sleep. Prevalence among Asian individuals did not change significantly in the 15-year period and was not significantly different from that of White individuals. Notably, when analyzed by age, the racial and ethnic disparities were greatest among young and middle-aged Black adults and slightly narrowed among the elderly.

This study expands the literature in several ways. First, we used data from 2004 to 2018 to describe trends in racial and ethnic disparities in sleep duration. Our findings regarding increasing prevalence of short sleep duration are consistent with previous NHIS studies,^11,12,31-33^ expanding them by quantifying the magnitude and significance of change in these racial and ethnic differences over the past 15 years and by analyzing disparities in long sleep duration. Of note, a study used data from the American Time Use Survey (ATUS) and found a slight increase in sleep duration from 2003 to 2016.^34^ Such a discrepancy may arise, in part, from how sleep duration is ascertained in each survey. In contrast with the holistic assessment of average sleep length in the NHIS, the ATUS is a phone-based survey that asks individuals to describe how they spent their day, starting at 4 a.m. the previous day and ending at 4 a.m. on the interview day.^35^ In addition, in the ATUS, self-reported time lying in bed may be recorded as sleep even if awake.^36^

Further research is needed to better understand this discrepancy. Second, we assessed the racial and ethnic differences in the association between each of these sleep duration outcomes and age. To the best of our knowledge, this has not been described before. Third, we stratified our findings by sex/gender and income, providing further insight in the characterization of these disparities. Fourth, we stratified by health status and found persistence in the disparities. Finally, we included Asian people in our analyses, finding that their estimates remained stable, without substantial differences with White individuals in 2018.

To understand why short sleep duration may be more common among Black people, it is important to discuss the influence of psychosocial stressors, such as race-based discrimination, on sleep health. The stress from perceived race-based discrimination (and its anticipation or vigilance) contributes to shorter sleep duration,^37^ and Black people in the US are more likely to experience this than other racial or ethnic groups. We showed that the disparity in short sleep duration remained stable over 15 years for Black people. Further, it has been reported that the effect of perceived discrimination on sleep duration is greater among Black women than among Black men,^38^ which could partially explain our finding that the short sleep racial gaps were the widest among Black women. Additional studies are needed to understand how these stressors derived from racial discrimination have changed over the last decades. Future work should also explore the extent to which Latino/Hispanic men may be facing increasing racial/ethnic-based discrimination or other social stressors that could explain their widening gap with White men over the study period.

Long sleep was also persistently more prevalent among Black individuals, particularly among Black women. This finding may be explained by persistent racial differences in prevalence and type of underlying health conditions and socioeconomic stressors that could potentially led to long sleep duration, including multimorbidity profiles (an indicator of multiple concurrent chronic conditions) and unemployment.^39-41^ Disparities in multimorbidity prevalence and unemployment rates persisted over the study period for Black individuals,^42,43^ which could support this explanation. However, further research is needed to understand these patterns and their causes.

The fact that the disparities were the widest among young and middle-aged adults may inform that factors related to working or employment conditions might be disproportionally preventing Black individuals from having adequate sleep.^44^ Notably, when analyzed over years and by age, the gap between Black and White individuals with low income was substantially narrower compared with the gap among those with middle or high income. This suggests that a higher income may prevent White people from experiencing sleep duration alterations but does not have such a protective association among Black people. This differential effect of income on sleep health is consistent with observations that higher educational attainment and professional responsibility is associated with lower odds of short sleep among White adults, and with greater odds among Black and Latino/Hispanic adults.^45-47^ It is possible that income in our study is an indicator of educational and professional attainment, and that Black individuals with higher income are more commonly exposed to stressors preventing adequate sleep, including higher racial discrimination.^48^

This study has several limitations. We relied on self-reported duration of sleep, which may be subject to recall and social desirability bias. Of note, across racial and ethnic groups, self-reported sleep has shown a low-to-moderate agreement with objective sleep duration measurement.^29,49-51^ When compared with polysomnography, White individuals overestimate their sleep duration by an average of 73 minutes whereas Black individuals overestimated it by 54 minutes.^29^ Such an overestimation may misclassify some participants’ sleep duration. Nonetheless, the potential 20-minute difference in self-reported sleep duration accuracy between White and Black individuals would only minimally explain the disparity between them, as suggested by our sensitivity analysis. Furthermore, self-reported sleep duration has important health implications, including consistent association with mortality across different populations^52-56^ and across racial/ethnic groups in the US.^57^ Additionally, for the entire study period, we lacked other information that may have provided a more in-depth understanding of these disparities in sleep health, including subjective sleep quality, efficiency, and timing.^58^ Lastly, it is possible that the declining NHIS response rates may have influenced our findings. Nonetheless, the NHIS design has several strategies to mitigate non-response bias (eMethods).

Our findings have important public health implications. These persistent disparities may contribute to other persistent racial and ethnic disparities in health. A study indicated that from 1999 to 2018, Black people had the highest prevalence of poor or fair health.^26^ Although the cross-sectional nature of these two studies prevents us from assessing causality, there is ample evidence that short or long sleep duration can cause detriments in health. Although the underlying cause of each sleep duration alteration may differ, both short and long sleep duration puts people at increased risk of depression, reduced quality of life, cardiovascular disease, diabetes, and death, among other conditions.^7,55,59-62^ Such a persistent disparity in sleep duration among Black people may thus be a cause and exacerbator of other health disparities, and may serve as an imperfect indicator of overall disparities in health and well-being. For the national objective of achieving health equity, understood as the assurance of the condition of optimal health for all people,^63^ it is thus instrumental to also strive for the elimination of socioeconomic and health conditions that prevent racial and ethnic minorities from achieving adequate sleep.

Our findings also have important implications for the design of public health interventions, suggesting that targeted efforts should be made to improve sleep health among Black and Latino/Hispanic individuals. The observed persistent—and growing—disparities in sleep duration serve as an additional indicator of the consequences of the artificial racial and ethnic hierarchy in which people of color encounter higher barriers to maintaining a healthy life, including income distribution inequality, racial segregation, restricted access to medical care, and exposure to social and environmental conditions that affect health and sleep such as light, noise, and air pollution. Thus, and as with other disparities, public policies may be ineffective in eliminating these racial and ethnic disparities in sleep without accounting for systemic racism as a fundamental cause.

In conclusion, from 2004 to 2018, there were significant differences in sleep duration by race and ethnicity, and the prevalence of unrecommended sleep duration was persistently higher among Black individuals. The disparities were highest for Black women, Black individuals who had middle or high income, and young and middle-aged Black adults. Given the importance of sleep to health, the prevalence of unrecommended sleep duration could be a contributor to health disparities.

## Supporting information

eMethods

## Data Availability

All data is publicly available from the Integrated Public Use Microdata Series Health Surveys (https://nhis.ipums.org/). The code used to analyze these data is publicly available at https://doi.org/10.5281/zenodo.6028375.

## DATA AVAILABILITY STATEMENT

Author César Caraballo had full access to all the data in the study and takes responsibility for the integrity of the data and the accuracy of the data analysis. All data is publicly available from the Integrated Public Use Microdata Series Health Surveys (https://nhis.ipums.org/). The code used to analyze these data is publicly available at https://doi.org/10.5281/zenodo.6028375.

