## Supplementary material for "Temporal Trends in Racial and Ethnic Disparities in Sleep Duration in the United States, 2004–2018": eMethods

### SUPPLEMENTAL MATERIAL

#### Trends in Racial and Ethnic Disparities in Sleep Duration in the United States: A National Health Interview Survey Analysis from 2004–2018

**eMethods.**

**eTable 1. Annualized Rate of Change in Short and Long Sleep Duration by Race and Ethnicity, 2004 to 2018.**

**eTable 2. Change in Short-Sleep Duration Prevalence by Race and Ethnicity and Health Status, 2004 to 2018.**

**eTable 3. Change in Long-Sleep Duration Prevalence by Race and Ethnicity and Health Status, 2004 to 2018.**

**eFigure 1. Study population flowchart.**

**eFigure 2. Unadjusted Sleep Duration Distribution by Race and Ethnicity.**

**eFigure 3. Short Sleep Sensitivity Analysis Accounting for Differences in Bias of Self-Reported Sleep Duration Between Black Individuals and White Individuals**

**eFigure 4. Adjusted Annual Prevalence of Short-Sleep Duration by Race and Ethnicity Among Those with Poor or Fair Health (A), And Among Those with Good to Excellent Health (B), 2004 to 2018.**

**eFigure 5. Adjusted Annual Prevalence of Long-Sleep Duration by Race and Ethnicity Among Those with Poor or Fair Health (A), And Among Those with Good to Excellent Health (B), 2004 to 2018.**

**eFigure 6. Racial and Ethnic Differences in the Relationship Between Age and Short-Sleep (A) and Long Sleep (B) Duration.**

**eFigure 7. Relationship Between Age and Short-Sleep Prevalence by Race/Ethnicity Stratified by Sex/Gender (A) and Stratified by Income Level (B).**

**eFigure 8. Relationship Between Age and Long Sleep Prevalence by Race/Ethnicity Stratified by Sex/Gender (A) and Stratified by Income Level (B).**

**eMethods.**

*About the National Health Interview Survey*

The National Health Interview Survey (NHIS) is a series of annual cross-sectional national surveys that provide information on the health of the noninstitutionalized population of the United States. The sample design uses a multistage area probability design, which adjusts for nonresponse and further allows for national representative sampling of households and individuals, including underrepresented groups. This survey consists of a questionnaire divided into 4 cores: Household Composition, Family Core, Sample Child Core, and Sample Adult Core. The Household Composition file collects basic and relationship information about all persons in a household. The Family Core file collects sociodemographic characteristics, basic indicators of health status, activity limitations, injuries, health insurance coverage, and access to and utilization of health care services. From each family, one sample child and one sample adult are randomly selected to gather more in-depth information for the Sample Child Core and Sample Adult Core, respectively. In our study, we used data from the Sample Adult Core files, complemented with demographic and socioeconomic characteristics.

*National Health Interview Survey Response Rates and Non-Response Bias Mitigation*

The NHIS assesses response rates at several different levels and reports two different sets of response rates, "Conditional" and "Final," each reported at the level of "family" and "adult." The difference between Conditional and Final rates is whether the household nonresponse is taken into account ("Final") or not ("Conditional"). [1]

The following are the response rates (both annual, and the pooled mean) for NHIS from 2004–2018:

| **Survey Year** | **Household Response Rate** | **Conditional Family Response Rate** | **Final Family Response Rate** | **Conditional Sample Adult Response Rate** | **Final Sample Adult Response Rate** |
|---|---|---|---|---|---|
| 2004 | 86.9 | 99.6 | 86.5 | 83.8 | 72.5 |
| 2005 | 86.5 | 99.5 | 86.1 | 80.1 | 69.0 |
| 2006 | 87.3 | 99.6 | 87.0 | 81.4 | 70.8 |
| 2007 | 87.1 | 99.4 | 86.6 | 78.3 | 67.8 |
| 2008 | 84.9 | 99.5 | 84.5 | 74.2 | 62.6 |
| 2009 | 82.2 | 99.3 | 81.6 | 80.1 | 65.4 |
| 2010 | 79.5 | 99.1 | 78.7 | 77.3 | 60.8 |
| 2011 | 82.0 | 99.2 | 81.3 | 81.6 | 66.3 |
| 2012 | 77.6 | 99.0 | 76.8 | 79.7 | 61.2 |
| 2013 | 75.7 | 99.0 | 74.9 | 81.7 | 61.2 |
| 2014 | 73.8 | 99.0 | 73.1 | 80.5 | 58.9 |
| 2015 | 70.1 | 98.9 | 69.3 | 79.7 | 55.2 |

| | | | | | |
|---|---|---|---|---|---|
| 2016 | 67.9 | 98.9 | 67.1 | 80.9 | 54.3 |
| 2017 | 66.5 | 98.9 | 65.7 | 80.7 | 53.0 |
| 2018 | 64.2 | 98.7 | 63.4 | 83.9 | 53.1 |
| Pooled Mean | 78.2 | 99.2 | 77.5 | 80.3 | 62.1 |

As such, the Conditional Response Rate for Sample Adults in our study period (2004–2018) is 80.3%, while the Final Response Rate for Sample Adults is 62.1%.

The NHIS design includes several adjustments to mitigate non-response bias.[2, 3] The Sample Adult weight, which we used in our analysis, includes design, ratio, non-response and post-stratification (adjustment of age-sex-race/ethnicity to Census population control totals). Of note, the sampling plan is redesigned after every decennial census to better measure the changing U.S. population. Thus, during the study period, the NHIS used weights derived from different Census-based population estimates: from 2000 Census for years 2004–2011, and from 2010 Census for years 2012-2018.

In addition, the NHIS uses oversamples Hispanic, Black, and Asian persons. This means that includes a larger sample of racial and ethnic minorities versus what would otherwise be required.[3] This increases the precision of the estimates obtained for these groups. For a more detailed description of the NHIS design please see Parsons et al, 2014.[4]

*Additional Description of Variables Included in the Study*

In the NHIS, hours of sleep responses are recoded into whole numbers, rounding values of 30 minutes or more up to the nearest hour or otherwise rounding down. The publicly available NHIS data includes data on 4 US regions: Northeast, North Central/Midwest, South, and West, based on where the housing unit of the survey participant was located. Health status is assessed on a 5-point scale (excellent, very good, good, fair, or poor) based on an individual's self-perceived general health. For this study, responses were dichotomized into poor or fair health status vs good to excellent health.

For descriptive purposes we classified categorical variables as follows: 3 categories for age (18–38, 40–64, ≥65 years); 2 categories for sex/gender; 2 categories for US citizenship status, 2 categories for family income (based on percent of family income relative to the federal poverty level from the Census Bureau: middle/high income

[≥200%] and low income [<200%]); 4 categories for education (more than bachelor's degree, some college, high school/general equivalency diploma, or less than high school); 2 categories for insurance coverage status at the time of the interview (insured and uninsured); 2 categories for marital status (married or living with partner, and not); 3 categories for employment status (with a job or working, not in labor force, and unemployed); 2 categories for flu vaccine in past 12 months (yes and no). We also included self-reported chronic conditions, including hypertension, diabetes, prior stroke or myocardial infarction, cancer, and emphysema or chronic bronchitis.

*Statistical Analysis*

To estimate the annual prevalence of short and long sleep duration prevalence rate for each racial/ethnic group we used separate multivariable multinomial logistic regression models. In these models, categorical sleep duration (short, recommended, and long) was the dependent variable, and age, a dummy variable for each region, and a dichotomous indicator for each year of interview were the independent variables. Age and region were centered on their overall mean for the study sample. The coefficients for each year, when combined with the intercept, then represented the logit of the annual rates of each sleep duration category adjusted for age and region. The results were used to generate estimated annual prevalence, using the inverse logit of each year effect as the annual prevalence and applying the method of parametric bootstrapping to calculate the standard error (SE) and the confidence interval (CI) for the transformed coefficients.[2]

To measure the racial and ethnic differences in short and long sleep duration, we subtracted the annual prevalence among White people from the annual prevalence among each of the other 3 groups for that year, also constructing SE for the differences. Then, to estimate trends over the study period, we used weighted linear regression models where the dependent variable was the adjusted annual sleep duration disturbance prevalence or difference, and the independent variable was time in years. To account for varying precision of each estimated prevalence or the difference over time, each observation was weighted by the inverse square of the SE.

To evaluate the association between race and ethnicity and each of these sleep duration disturbances by age, we used a similar approach as above. We used multinomial regression model with categorical sleep duration as the dependent variable and age groups as the independent variables. Age was categorized by 5-year groups for this analysis. To measure the racial and ethnic differences in short and long with age, we subtracted the age group prevalence among White people from the age group prevalence among each of the other 3 groups for that year, also constructing SE for the differences.

To estimate the annual low-income prevalence by race and ethnicity we used the mean annual estimate obtained by separate multinomial logistic regressions using a similar approach as above, but with each of the multiply imputed low-income variables as the dependent variable and an indicator for each year as the independent variables. Similarly, we used the mean prevalence estimate of each sleep duration outcome (short or long) obtained from separate multinomial regressions using each of the income groups.[3]

Lastly, we performed a sensitivity analysis to assess if the observed disparities in short sleep duration were explained by differences in self-reported sleep duration bias. In this sensitivity analysis, considering that the publicly available NHIS data includes sleep duration in integers, we artificially created hourly subdeciles by randomly and equally distributing individuals into 10 bins based on their NHIS sleep duration value (e.g., *X* number of individuals that reported 6 hours were randomly assigned to equally distributed deciles of an hour between 6 and 7 hours; each bin containing with *X*/10 number of individuals). We then added 73 minutes and 54 minutes to White and Black individuals' sleep duration, respectively, based on the mean sleep duration overestimation of each race group when compared with polysomnography.[7] We then performed the same multinomial logistic regressions as above to assess the annual prevalence of short, recommended, and long sleep duration for each race; and the differences between both groups.

**eTable 1. Annualized Rate of Change in Short and Long Sleep Duration by Race and Ethnicity, 2004 to 2018.**

| | **Asian individuals** | **Black individuals** | **Latino/Hispanic individuals** | **White individuals** |
|---|---|---|---|---|
| | Percentage points (95% CI), p value | Percentage points (95% CI), p value | Percentage points (95% CI), p value | Percentage points (95% CI), p value |
| **Short sleep duration (<7 hours)** | | | | |
| Overall | +0.21 (-0.03 to 0.44), 0.08 | +0.54 (0.38 to 0.69), <0.001 | +0.55 (0.39 to 0.72), <0.001 | +0.27 (0.16 to 0.38), <0.001 |
| *Women* | +0.33 (0.07 to 0.59), 0.02 | +0.60 (0.42 to 0.78), <0.001 | +0.57 (0.37 to 0.76), <0.001 | +0.31 (0.20 to 0.42), <0.001 |
| *Men* | +0.09 (-0.25 to 0.44), 0.57 | +0.46 (0.24 to 0.68), 0.001 | +0.52 (0.30 to 0.75), <0.001 | +0.23 (0.10 to 0.37), 0.003 |
| *Low income* | +0.45 (0.08 to 0.83), 0.02 | +0.69 (0.51 to 0.87), <0.001 | +0.61 (0.42 to 0.79), <0.001 | +0.38 (0.21 to 0.56), <0.001 |
| *Mid/high income* | +0.14 (-0.17 to 0.45), 0.35 | +0.45 (0.23 to 0.66), 0.001 | +0.49 (0.32 to 0.65), <0.001 | +0.25 (0.14 to 0.35), <0.001 |
| **Long Sleep Duration (>9 hours)** | | | | |
| Overall | -0.04 (-0. 10 to 0.03), 0.28 | -0.09 (-0.16 to -0.03), 0.005 | -0.06 (-0.12 to 0.01), 0.08 | -0.01 (-0.06 to 0.03), 0.59 |
| *Women* | -0.06 (-0.13 to 0.02), 0.13 | -0.05 (-0.11 to 0.01), 0.10 | -0.10 (-0.16 to -0.04), 0.003 | +0.01 (-0.04 to 0.05), 0.78 |
| *Men* | -0.02 (-0.12 to 0.08), 0.67 | -0.16 (-0.28 to -0.05), 0.007 | -0.02 (-0.11 to 0.06), 0.54 | -0.03 (-0.09 to 0.02), 0.24 |
| *Low income* | -0.06 (-0.20 to 0.08), 0.37 | -0.20 (-0.30 to -0.10), 0.001 | -0.08 (-0.18 to 0.01), 0.09 | +0.01 (-0.10 to 0.11), 0.90 |
| *Mid/high income* | -0.01 (-0.09 to 0.06), 0.70 | -0.02 (-0.08 to 0.04), 0.48 | -0.01 (-0.09 to 0.08), 0.90 | -0.01 (-0.04 to 0.02), 0.36 |

Data source is the National Health Interview Survey from years 2004 to 2018. For change in prevalence: a positive sign (+) means the prevalence and a negative sign (-) means it decreased. Prevalence estimates were adjusted by age and region.
Abbreviations: CI, confidence interval

**eTable 2. Change in Short-Sleep Duration Prevalence by Race and Ethnicity and Health Status, 2004 to 2018.**

| | Asian | Black | Latino/Hispanic | White |
|---|---|---|---|---|
| **Short-sleep duration** | Percentage points (95% CI), p value | Percentage points (95% CI), p value | Percentage points (95% CI), p value | Percentage points (95% CI), p value |
| **Annualized rate of change in prevalence** | | | | |
| Poor or fair health | +0.19 (-0.70 to 1.09), 0.65 | +0.53 (0.27 to 0.78), 0.001 | +0.67 (0.29 to 1.05), 0.002 | +0.42 (0.26 to 0.60), <0.001 |
| Good to excellent health | +0.21 (-0.04 to 0.46), 0.09 | +0.56 (0.39 to 0.72), <0.001 | +0.53 (0.37 to 0.69), <0.001 | +0.25 (0.14 to 0.37), <0.001 |
| **Absolute change in prevalence, 2004–2018** | | | | |
| Poor or fair health | +4.28 (-12.63 to 21.19), 0.62 | +10.32 (3.76 to 16.88), 0.002 | +13.64 (6.22 to 21.06), <0.001 | +3.08 (-0.38 to 6.54), 0.08 |
| Good to excellent health | +1.37 (-3.41 to 6.15), 0.58 | +5.90 (2.47 to 9.32), <0.001 | +5.51 (2.74 to 8.28), <0.001 | +3.21 (2.00 to 4.43), <0.001 |
| **Difference with White, 2004** | | | | |
| Poor or fair health | +0.39 (-13.38 to 14.16), 0.96 | +3.47 (-1.40 to 8.35), 0.16 | -4.49 (-9.36 to 0.38), 0.07 | - |
| Good to excellent health | +4.19 (0.64 to 7.74), 0.02 | 7.17 (4.90, 9.44), <0.001 | -1.19 (-3.10 to 0.73), 0.23 | - |
| **Difference with White, 2018** | | | | |
| Poor or fair health | +1.59 (-8.82 to 12.00), 0.77 | +10.71 (5.13 to 16.30), <0.001 | +6.06 (-0.52 to 12.64), 0.07 | - |
| Good to excellent health | +2.34 (-1.08 to 5.77), 0.18 | +9.85 (7.02 to 12.69), <0.001 | +1.11 (-1.23 to 3.45), 0.35 | - |
| **Absolute change in difference with White, 2004–2018** | | | | |
| Poor or fair health | +1.20 (-16.06 to 18.46), 0.89 | +7.24 (-0.18 to 14.65), 0.06 | +10.56 (2.37 to 18.74), 0.01 | - |
| Good to excellent health | -1.85 (-6.78 to 3.09), 0.46 | +2.68 (-0.95 to 6.32), 0.15 | +2.29 (-0.73 to 5.32), 0.14 | - |

Legend: Data source is the National Health Interview Survey from years 2004 to 2018. For change in prevalence and change in difference: a positive sign (+) means the prevalence of short sleep (or its difference with White people) increased and a negative sign (-) means it decreased. Short-sleep duration prevalence was adjusted by age and region.
Abbreviations: CI, confidence interval

**eTable 3. Change in Long-Sleep Duration Prevalence by Race and Ethnicity and Health Status, 2004 to 2018.**

| | Asian | Black | Latino/Hispanic | White |
|---|---|---|---|---|
| **Long-sleep duration** | Percentage points (95% CI), p value | Percentage points (95% CI), p value | Percentage points (95% CI), p value | Percentage points (95% CI), p value |
| **Annualized rate of change in prevalence** | | | | |
| Poor or fair health | +0.08 (-0.12 to 0.27), 0.42 | -0.05 (-0.18 to 0.08), 0.42 | -0.11 (-0.27 to 0.05), 0.15 | -0.03 (-0.13 to 0.08), 0.62 |
| Good to excellent health | -0.06 (-0.12, 0.00), 0.06 | -0.08 (-0.14 to -0.02), 0.02 | -0.04 (-0.09 to 0.02), 0.14 | 0.00 (-0.04 to 0.04), 0.89 |
| **Absolute change in prevalence, 2004–2018** | | | | |
| Poor or fair health | +2.69 (-4.13 to 9.50), 0.44 | +1.21 (-1.92 to 4.39), 0.46 | -4.84 (-8.02 to -1.65), 0.003 | +0.50 (-1.39 to 2.39), 0.60 |
| Good to excellent health | -0.71 (-2.33 to 0.91), 0.39 | -1.69 (-3.27 to -0.11), 0.04 | -0.63 (-1.64 to 0.39), 0.23 | +0.29 (-0.17 to 0.75), 0.22 |
| **Difference with White, 2004** | | | | |
| Poor or fair health | -4.78 (-9.73 to -0.18), 0.06 | -1.71 (-3.92 to 0.49), 0.13 | 0.00 (-2.67 to 2.68), 0.99 | - |
| Good to excellent health | -0.35 (-1.78 to 1.09), 0.64 | +3.25 (1.98 to 4.52), <0.001 | +0.54 (-0.17 to 1.25), 0.14 | - |
| **Difference with White, 2018** | | | | |
| Poor or fair health | -2.59 (-7.63 to 2.45), 0.31 | -1.01 (-3.98 to 1.97), 0.51 | -5.33 (-7.89 to -2.77), <0.001 | - |
| Good to excellent health | -1.34 (-2.22, -0.46), 0.003 | +1.28 (0.23, 2.33), 0.02 | -0.38 (-1.23 to 0.48), 0.39 | - |
| **Absolute change in difference with White, 2004–2018** | | | | |
| Poor or fair health | +2.18 (-4.89 to 9.25), 0.55 | +0.71 (-2.99 to 4.41), 0.71 | -5.34 (-9.04 to -1.64), 0.005 | - |
| Good to excellent health | -1.00 (-2.68 to 0.69), 0.25 | -1.98 (-3.62 to -0.33), 0.02 | -0.91 (-2.03 to 0.20), 0.11 | - |

Legend: Data source is the National Health Interview Survey from years 2004 to 2018. For change in prevalence and change in difference: a positive sign (+) means the prevalence of long sleep (or its difference with White people) increased and a negative sign (-) means it decreased. Long-sleep duration prevalence was adjusted by age, sex, and region.
Abbreviations: CI, confidence interval

**eFigure 1. Study Population Flowchart.**

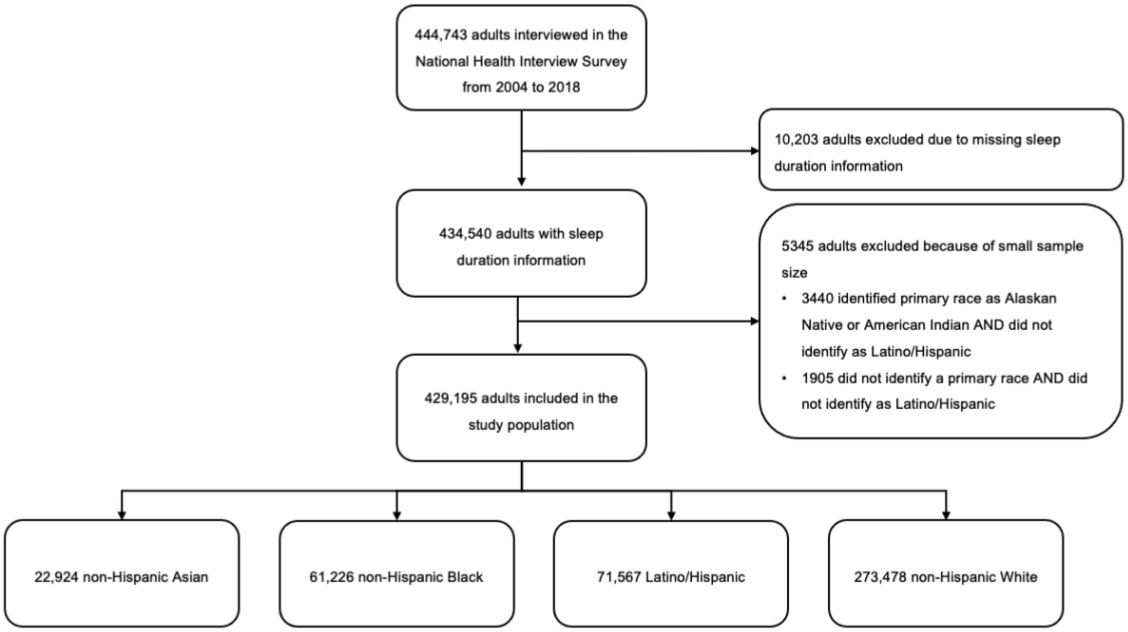


Legend: The mutually exclusive racial and ethnic groups were built based on participants' self-reported selection of primary race and ethnicity.

**eFigure 2. Unadjusted Sleep Duration Distribution by Race and Ethnicity.**

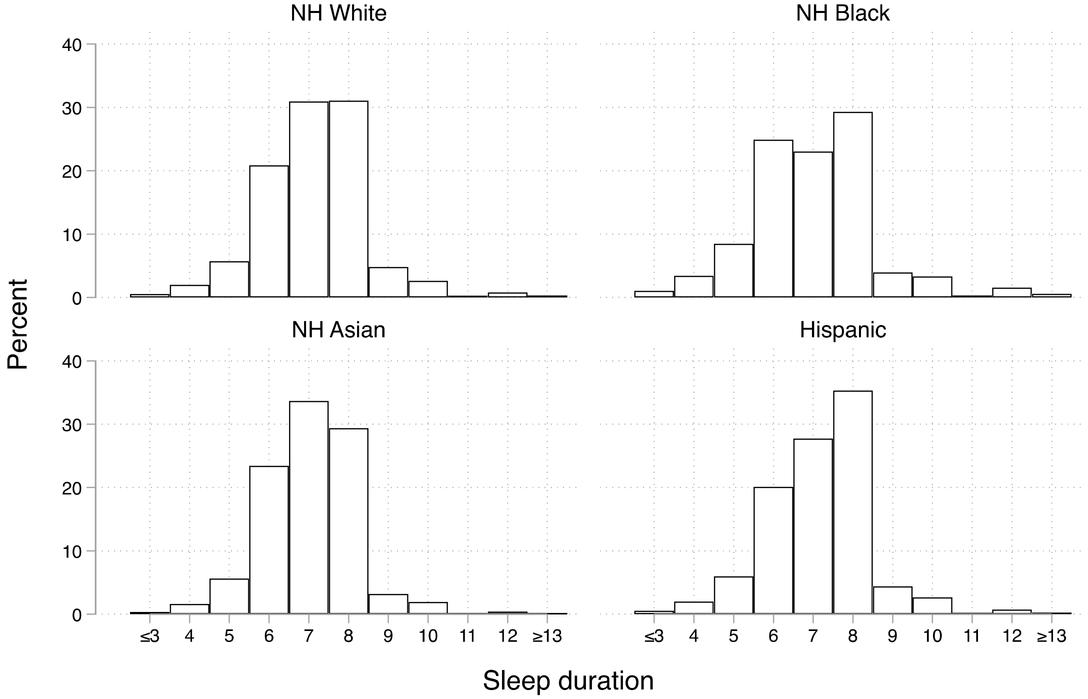


Legend: Data source is the National Health Interview Survey from years 2004 to 2018. All percentages are unweighted and unadjusted. NH, non-Hispanic.

**eFigure 3. Short Sleep Sensitivity Analysis Accounting for Differences in Bias of Self-Reported Sleep Duration Between Black Individuals and White Individuals**

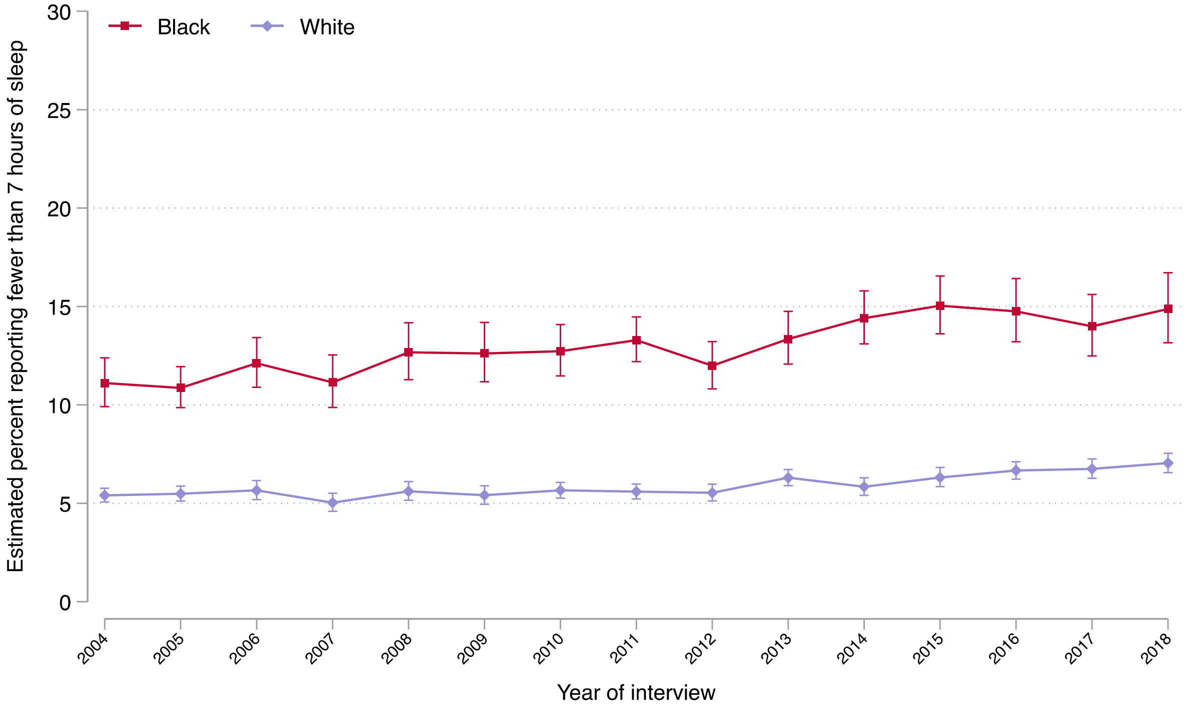


| | Main Analysis | | | | Sensitivity Analysis | | | |
|---|---|---|---|---|---|---|---|---|
| | **Black individuals** Percentage points (95% CI) | **P value** | **White individuals** Percentage points (95% CI) | **P value** | **Black individuals** Percentage points (95% CI) | **P value** | **White individuals** Percentage points (95% CI) | **P value** |
| *Absolute change in prevalence, 2004–2018* | +6.39 (3.32 to 9.46) | <0.001 | +3.22 (2.06 to 4.38) | <0.001 | +3.76 (+1.59 to +5.93) | <0.001 | +1.64 (+1.04 to +2.24) | <0.001 |
| *Difference with White, 2004* | +7.51 (5.45 to 9.57) | <0.001 | - | - | +5.71 (+4.41 to +7.00) | <0.001 | - | - |
| *Difference with White, 2018* | +10.68 (8.12 to 13.24) | <0.001 | - | - | +7.83 (+5.99 to +9.67) | <0.001 | - | - |
| *Absolute change in difference with White, 2004–2018* | +3.17 (-0.11 to 6.46) | 0.06 | - | - | +2.12 (-0.13 to +4.37) | 0.07 | - | - |

Data source is the National Health Interview Survey from years 2004 to 2018. Short-sleep duration was defined as fewer than 7 (<7) hours of sleep in a 24-hour period. For change in prevalence and change in difference: a positive sign (+) means the prevalence (or its difference with White individuals) increased and a negative sign (-) means it decreased. Prevalence estimates were adjusted by age and region. See details of this sensitivity analysis in eMethods.

Abbreviations: CI, confidence interval

**eFigure 4. Adjusted Annual Prevalence of Short-Sleep Duration by Race and Ethnicity Among Those with Poor or Fair Health (A), And Among Those with Good to Excellent Health (B), 2004 to 2018.**

**A)**

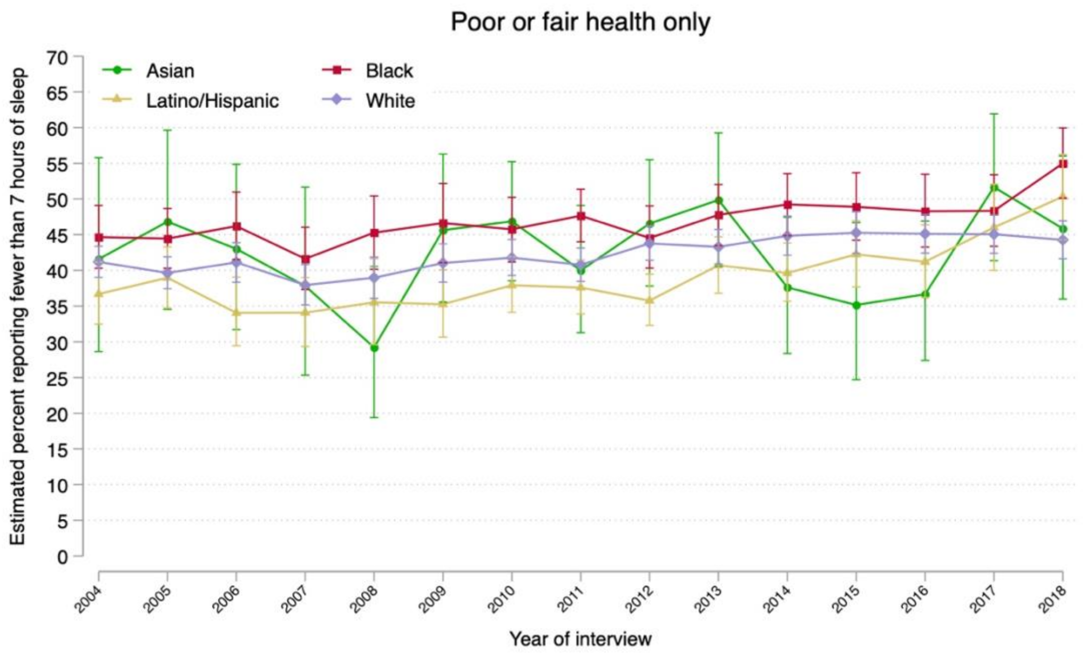


**B)**

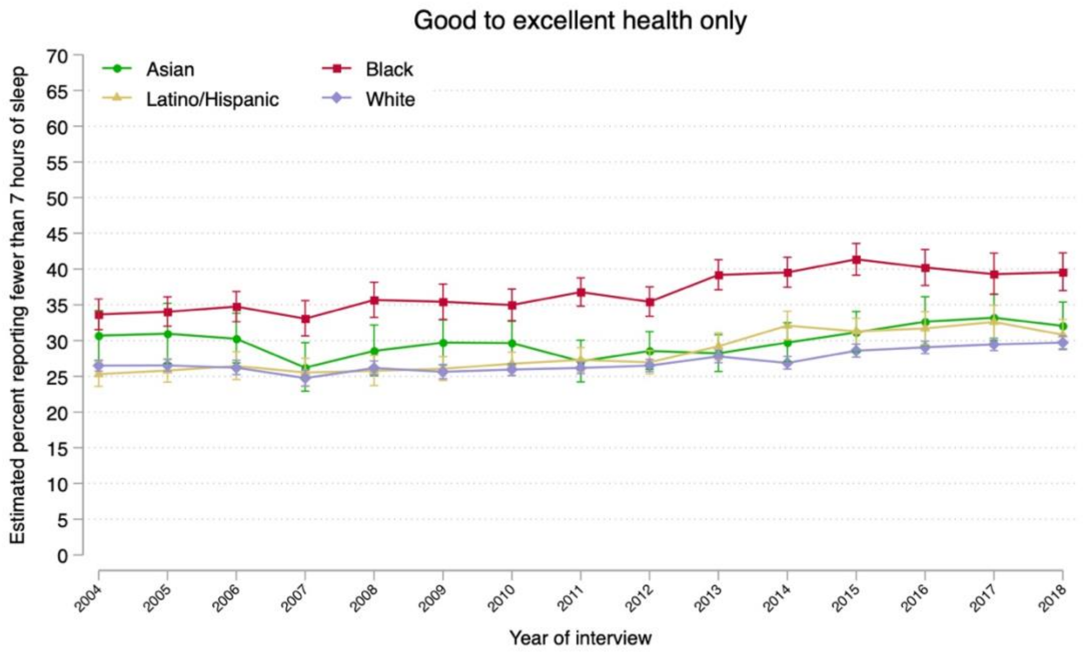

**eFigure 5. Adjusted Annual Prevalence of Long-Sleep Duration by Race and Ethnicity Among Those with Poor or Fair Health (A), And Among Those with Good to Excellent Health (B), 2004 to 2018.**

**A)**

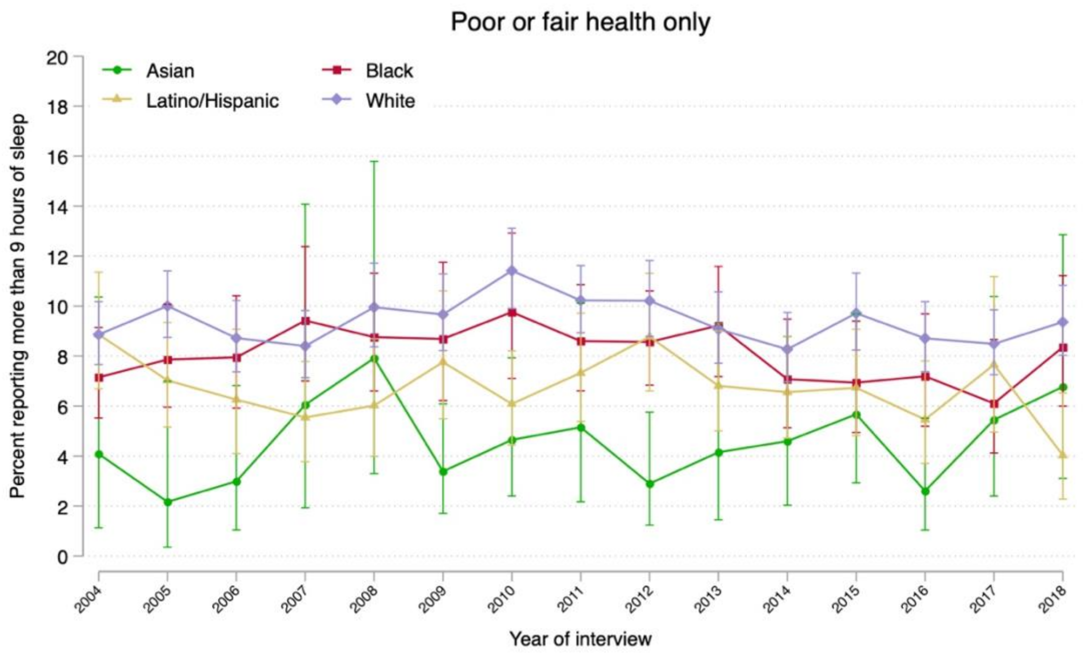


**B)**

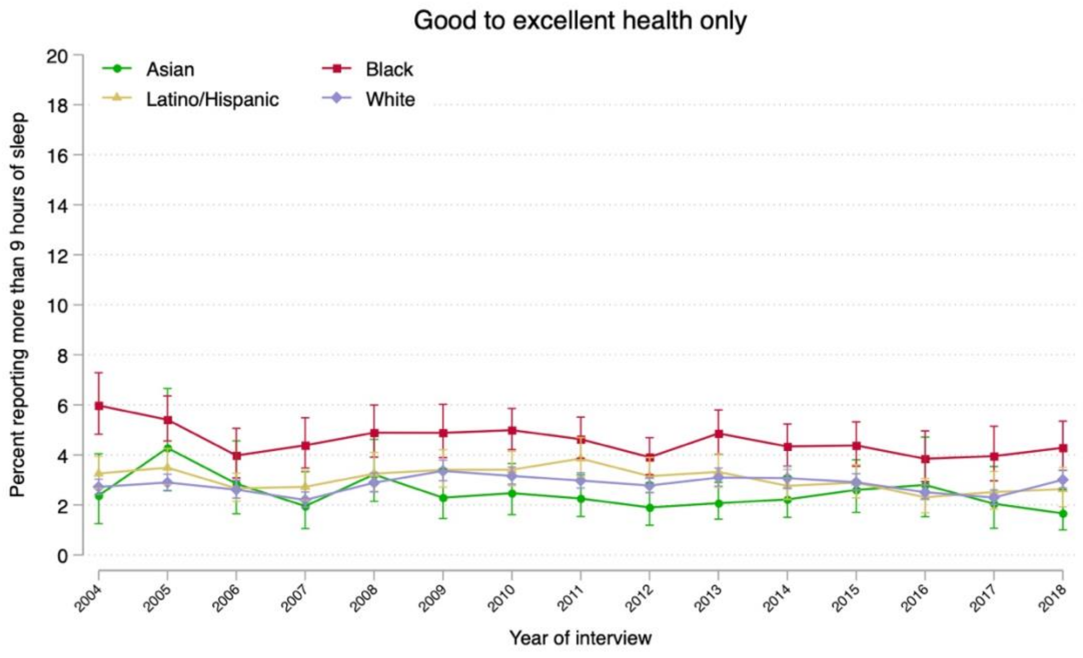

**eFigure 6. Racial and Ethnic Differences in the Relationship Between Age and Short-Sleep (A) and Long Sleep (B) Duration**

**A)**

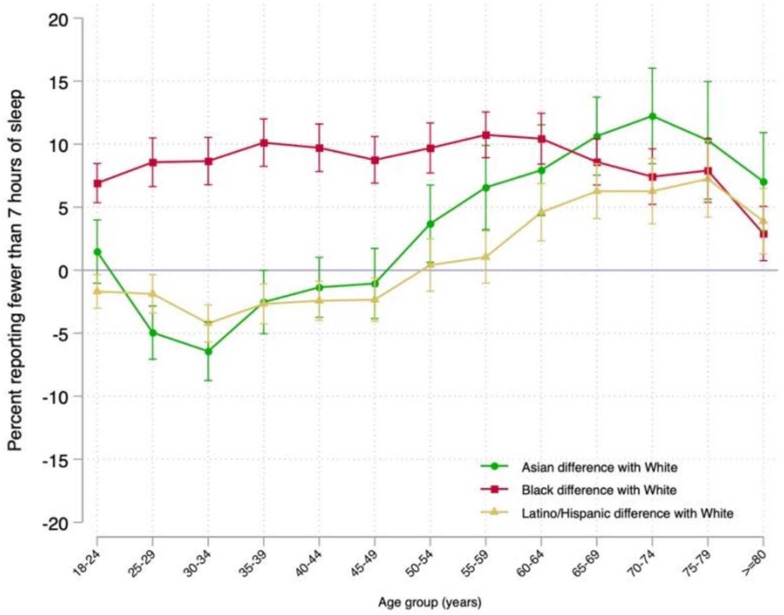


**B)**

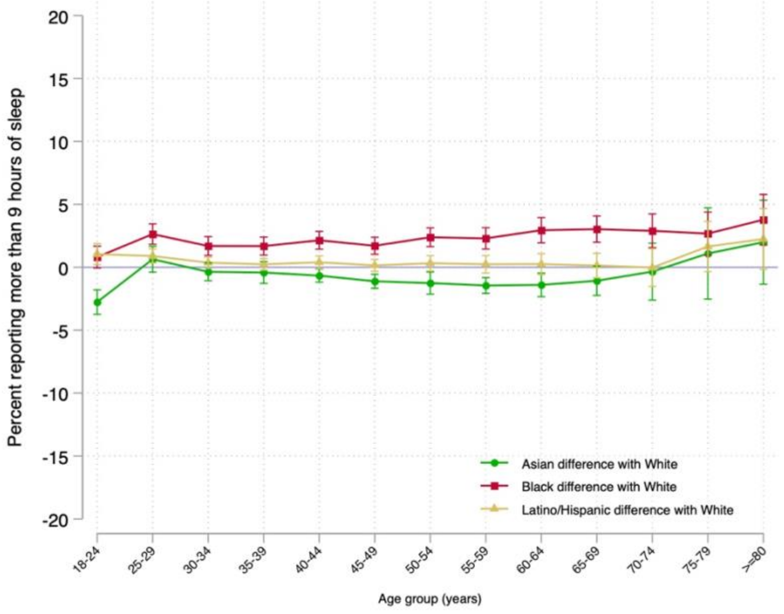

**eFigure 7. Relationship Between Age and Short-Sleep Prevalence by Race/Ethnicity Stratified by Sex/Gender (A) and Stratified by Income Level (B).**

**A)**

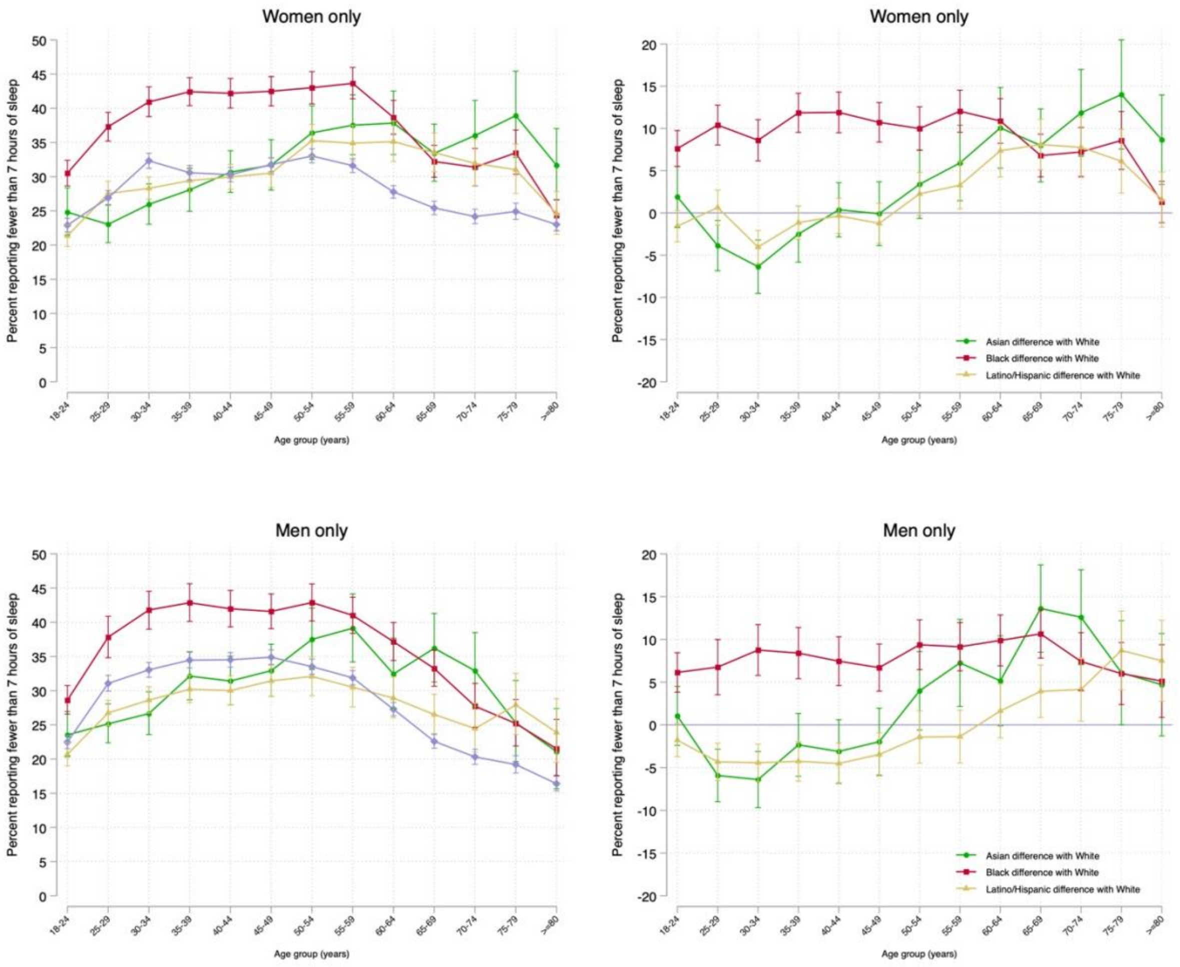

**B)**

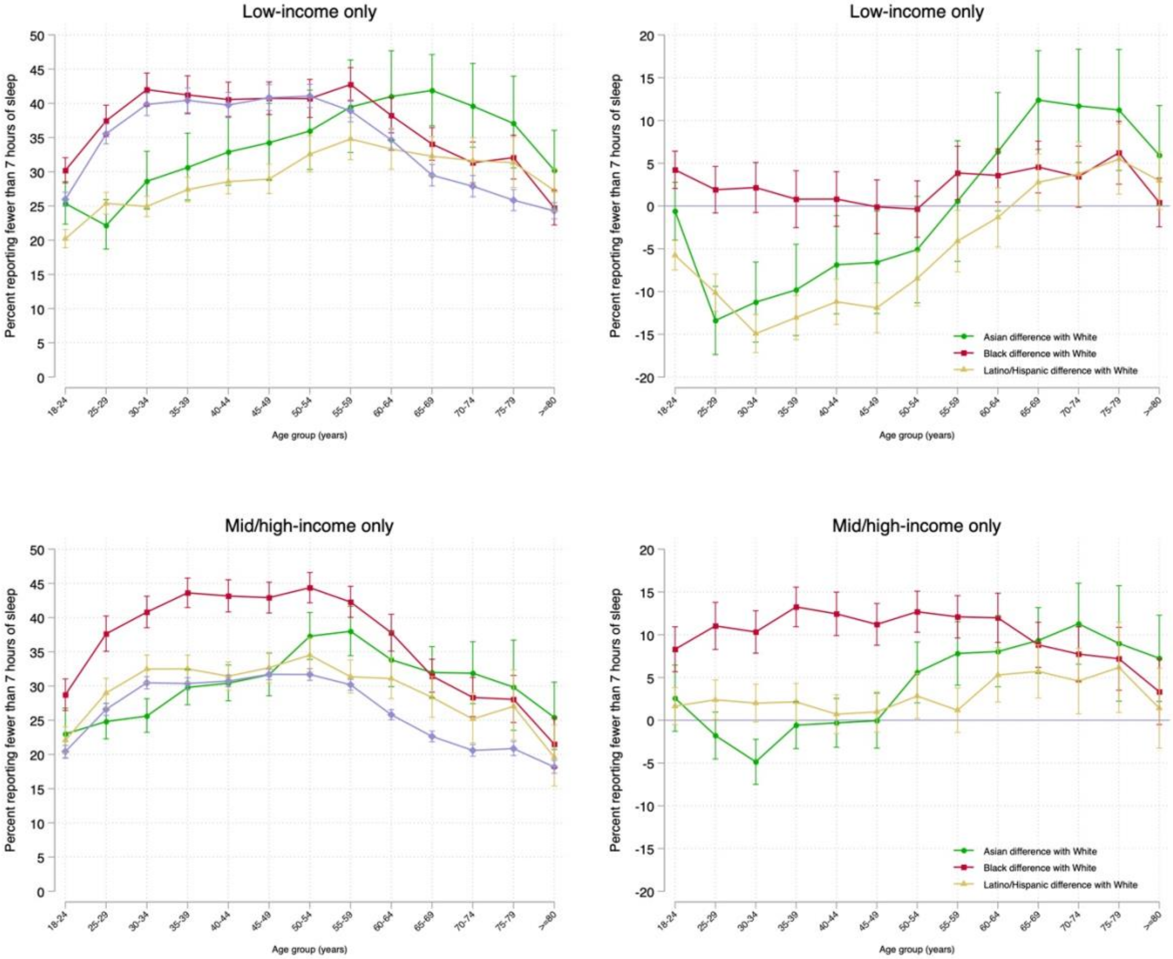

Low-income only
Percent reporting fewer than 7 hours of sleep
Age group (years)
18-24
25-29
30-34
35-39
40-44
45-49
50-54
55-59
60-64
65-69
70-74
75-79
>=80
Low-income only
Asian difference with White
Black difference with White
Latino/Hispanic difference with White
Mid/high-income only
Mid/high-income only

**eFigure 8. Relationship Between Age and Long-Sleep Prevalence by Race/Ethnicity Stratified by Sex/Gender (A) and Stratified by Income Level (B).**

**A)**

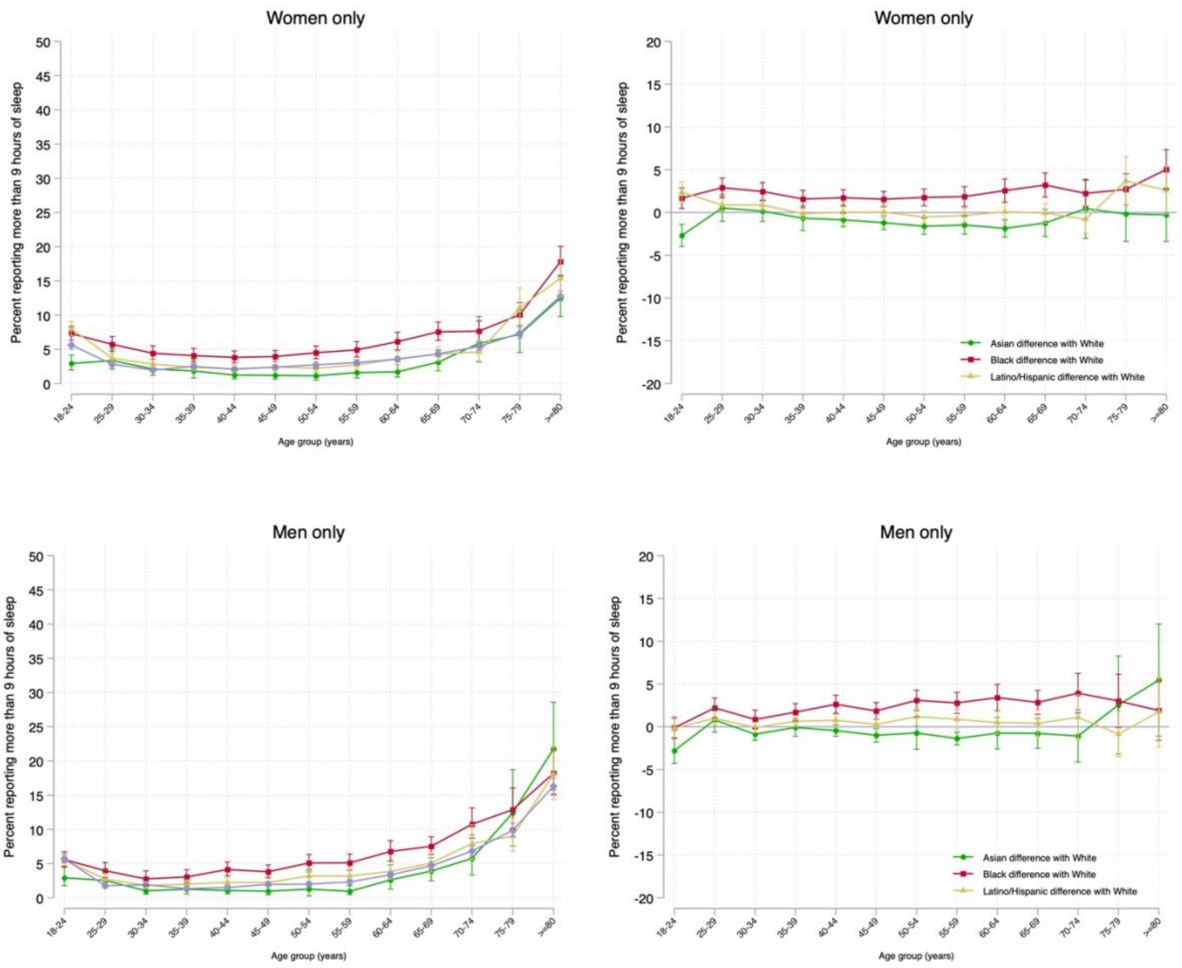

**B)**

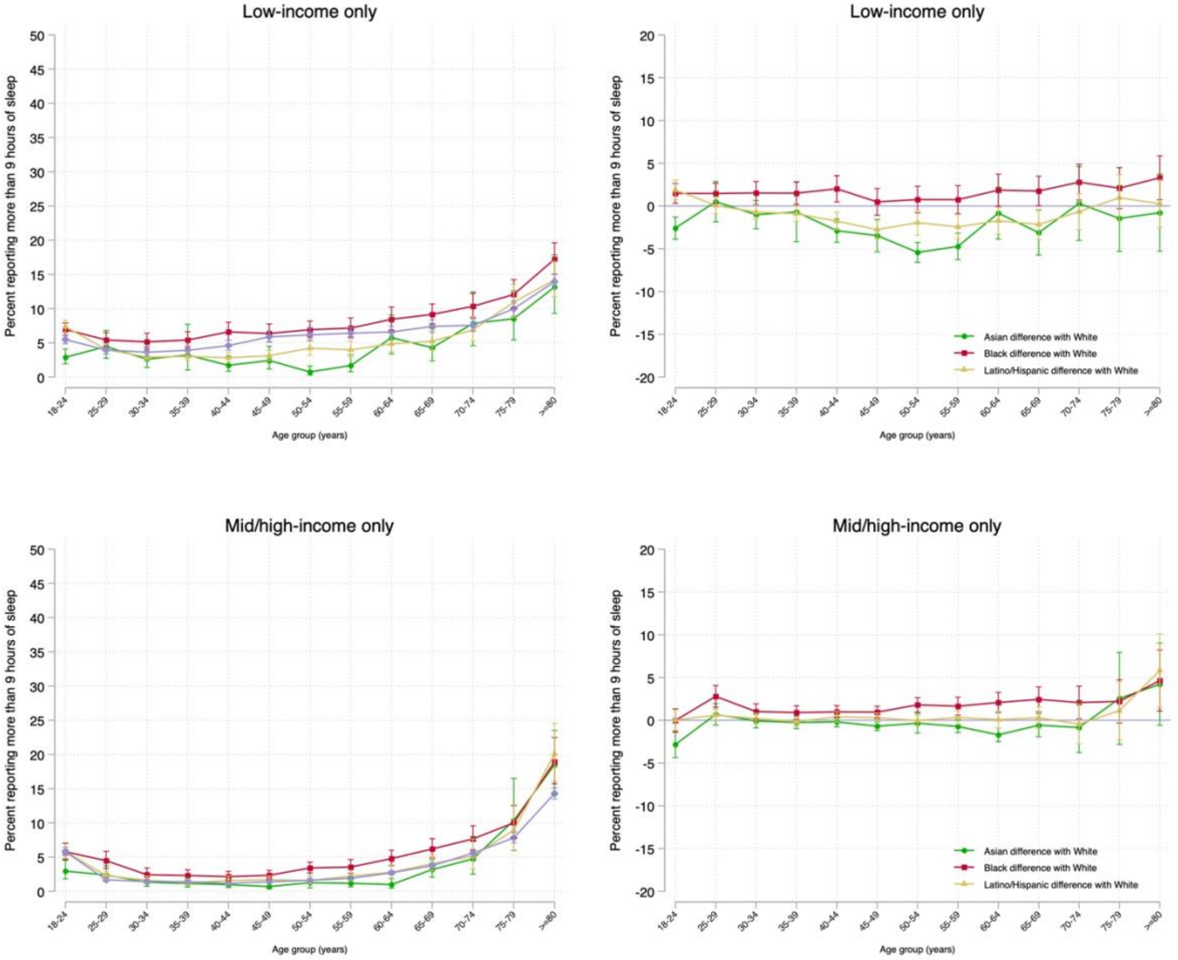

Low-income only
Percent reporting more than 9 hours of sleep
Age group (years)
18-24
25-29
30-34
35-39
40-44
45-49
50-54
55-59
60-64
65-69
70-74
75-79
>=80
Low-income only
Asian difference with White
Black difference with White
Latino/Hispanic difference with White
Mid/high-income only
Mid/high-income only
